# Safety and immunogenicity of SARS-CoV-2 self-amplifying RNA vaccine expressing anchored RBD: a randomised, observer-blind, phase 1 study

**DOI:** 10.1101/2022.11.21.22281000

**Authors:** Wataru Akahata, Takashi Sekida, Takuto Nogimori, Hirotaka Ode, Tomokazu Tamura, Kaoru Kono, Yoko Kazami, Ayaka Washizaki, Yuji Masuta, Rigel Suzuki, Kenta Matsuda, Mai Komori, Amber Morey, Keiko Ishimoto, Misako Nakata, Tomoko Hasunuma, Takasuke Fukuhara, Yasumasa Iwatani, Takuya Yamamoto, Jonathan F Smith, Nobuaki Sato

**Affiliations:** VLP Therapeutics Japan, LLC., Marunouchi, Chiyoda-ku, Tokyo, Japan; Clinical Research Center, National Hospital Organization Nagoya Medical Center, Nagoya, Aichi, Japan; Department of Microbiology and Immunology, Faculty of Medicine, Hokkaido University, Sapporo, Japan; Laboratory of Immunosenescence, Center for Vaccine and Adjuvant Research, National Institutes of Biomedical Innovation, Health and Nutrition, Osaka, Japan; VLP Therapeutics, Inc., Gaithersburg, MD, USA; Department of Research, Kitasato University, Kitasato Institute Hospital, Tokyo, Japan; Laboratory of Virus Control, Research Institute for Microbial Diseases, Osaka University, Suita, Japan; Division of Basic Medicine, Nagoya University Graduate School of Medicine, Nagoya, Japan

## Abstract

**BACKGROUND:** VLPCOV-01 is a lipid nanoparticle-encapsulated self-amplifying RNA (saRNA) vaccine that expresses a membrane-anchored receptor-binding domain (RBD) derived from the SARS-CoV-2 spike protein.

**METHODS:** A phase 1 study of VLPCOV-01 was conducted at Medical Corporation Heishinkai OPHAC Hospital, Japan. Participants aged 18 to 55 or ≥65 years who had completed two doses of the BNT162b2 mRNA vaccine 6 to 12 months previously were randomised to receive one intramuscular vaccination of 0·3, 1·0, or 3·0 μg VLPCOV-01, 30 μg BNT162b2, or placebo between February 16, 2022, and March 17, 2022. Solicited adverse events were collected up to 6 days post-administration. Interim immunogenicity analyses included SARS-CoV-2 IgG and neutralising antibody titres. Follow-up for safety and immunogenicity evaluation is ongoing. (The trial is registered: jRCT2051210164).

**FINDINGS:** 92 healthy adults were enrolled, with 60 participants receiving VLPCOV-01. No serious adverse events were reported up to 26 weeks, and no prespecified trial-halting events were met. VLPCOV-01 induced robust IgG titres against SARS-CoV-2 RBD protein that were maintained up to 26 weeks in non-elderly participants, with geometric means ranging from 5037 (95% CI 1272–19,940) at 0·3 μg to 12,873 (95% CI 937–17,686) at 3 μg, in comparison to 3166 (95% CI 1619–6191) with 30 μg BNT162b2. Among elderly participants, IgG titres at 26 weeks post-vaccination with 3 μg VLPCOV-01 were 9865 (95% CI 4396–22138) compared to 4183 (95% CI 1436–12180) following vaccination with 30 μg BNT162b2. Pseudovirus neutralising antibody responses were observed against multiple SARS-CoV-2 variants and strongly correlated with anti-SARS-CoV-2 IgG (*r*=0·950, p<0·001).

**INTERPRETATION:** VLPCOV-01 is immunogenic following low dose administration, with anti-SARS-CoV-2 immune responses comparable to BNT162b2. These findings support further development of VLPCOV-01 as a COVID-19 booster vaccine and the potential for saRNA vectors as a vaccine platform.

**FUNDING:** Supported by AMED, Grant No. JP21nf0101627.

## Introduction

The coronavirus disease 2019 (COVID-19) pandemic, caused by severe acute respiratory syndrome coronavirus 2 (SARS-CoV-2), has had a significant global impact, with over 629 million cases and more than 6·5 million deaths reported as of November 7, 2022.^1^ In response to the emergence of SARS-CoV-2 in Wuhan, China in December 2019, there was rapid clinical development of SARS-CoV-2 vaccines, with >300 vaccines in development as of May 2022. The World Health Organization (WHO) has approved ten COVID-19 vaccines for global use, reflecting eight distinct vaccine products and four distinct vaccine platforms.^2^ Widespread rollout of these vaccines has saved tens of millions of lives.^1,3^ Despite the availability of vaccines that have demonstrated efficacy against COVID-19,^4-10^ the rapidly changing epidemiology, emergence of new variants, and increased transmissibility despite previous COVID-19 vaccination necessitate new strategies for prevention.^2^

Self-amplifying RNA (saRNA) encapsulated in lipid nanoparticles (LNPs) is a novel technology for use in vaccines.^11^ Vaccines developed using saRNA are promising candidates for meeting the challenges of pandemics due to unique features, including low dose administration and a readily modifiable antigenic domain, enabling rapid development of vaccines in response to emerging variants of concern.^11,12^ We developed a novel LNP-encapsulated saRNA COVID-19 vaccine (VLPCOV-01) expressing a membrane-anchored receptor-binding domain (RBD) that utilises an alphavirus RNA amplification system.^13^ The self-amplification process results in efficient expression of the gene of interest and a longer duration of expression than that which is observed with the mRNA vaccine platforms.^12^ VLPCOV-01 encodes replicase and transcriptase functions composed of the alphavirus nonstructural proteins (nsP1-4) from the attenuated TC-83 strain of Venezuelan equine encephalitis virus (VEEV) as well as the RBD of the spike protein from SARS-CoV-2. The RBD sequence is fused with a C-terminal transmembrane domain (RBD-TM), focusing the immune response on the RBD that contains most of the neutralising epitopes.^14^ Preclinical evaluation of VLPCOV-01 in hamster and non-human primate models using LNP delivery systems demonstrated efficient induction of T-cell and B-cell responses that conferred protection against SARS-CoV-2 challenges. Furthermore, RBD-specific antibodies against variants of concern, including Delta and Omicron variants, were maintained for at least 6 months in non-human primates (*in submission*).

Here, we present the results of a phase 1 study of VLPCOV-01 in healthy adults who had already received primary vaccination with BNT162b2 to confirm the safety and efficacy of VLPCOV-01 as a booster vaccine.

## Methods

### Study design and participants

We assessed the safety, immunogenicity, and dosage of a single booster dose of VLPCOV-01 in a randomised, single-centre, placebo- and active-controlled, observer-blind phase 1 study. The study protocol was reviewed and approved by the Medical Corporation Heishinkai OPHAC Hospital institutional review board, and the trial was conducted at Medical Corporation Heishinkai OPHAC Hospital. No important changes to the methods were made following trial commencement.

The planned number of participants as set out in the study protocol was 92. This was deemed an appropriate target to evaluate the immunogenicity and safety of the investigational drug. Eligible participants were healthy Japanese adults aged 18 to 55 or ≥65 years who had completed two doses of the mRNA vaccine BNT162b2 6 to 12 months previously. Key exclusion criteria were a history of COVID-19, pregnant and lactating females, a history or presence of a serious cardiovascular, haematological, respiratory, hepatic, renal, gastrointestinal, and/or neuropsychiatric disease, and previously received or planned to receive treatment with any drug or therapy considered to affect the immunogenicity assessments. All participants provided written informed consent before enrolment. Full eligibility criteria are described in the study protocol.

### Randomisation and masking

The principal investigator appointed a study drug randomization supervisor who prepared the randomization schedule prior to participant enrolment and was responsible for storing and managing it until study unblinding. Potential participants were assigned a screening number by the principal or sub investigator, assessed for suitability to participate in the study, and upon completion of screening, were allocated a participant identification code. Participants were stratified into two age subgroups: 18 to 55 years (non-elderly cohort) and ≥65 years (elderly cohort). Participants in each group were randomised to receive VLPCOV-01 at doses of 0·3, 1·0, or 3·0 μg, 30-μg BNT162b2, or placebo. Transition to the next VLPCOV-01 dose cohort in the same age group, and from the non-elderly cohort to the elderly cohort, was performed after the principal investigator determined that there were no medical concerns based on the results of the tests and examinations performed up to day 4 for each cohort. Assessors and participants were blinded for the study. The principal investigator assigned unblinded medical staff in advance who allocated, prepared and administered the study drugs and were not involved in the assessments of participants after administration.

### Procedures

The investigational vaccine, VLPCOV-01, was compared to 30 μg BNT162b2 and placebo (0·9% saline). VLPCOV-01, developed by VLP Therapeutics, is a saRNA vaccine against COVID-19, stabilised by and delivered with LNPs. The saRNA expresses the nonstructural proteins of the alphavirus from the 5’ end of the RNA, as well as an engineered RBD sequence of SARS-CoV-2 (Wuhan stain; GenBank Yp_009724390.1) from the subgenomic promoter, thus allowing amplification of the replicon RNA in transfected cells and high-level expression of the RBD construct. All study drugs were stored in accordance with study drug management procedures.

The study drug was administered as a single dose by the intramuscular route into the deltoid region of the upper arm. Participants were carefully monitored for ≥30 minutes after administration for assessment of reactogenicity. Medical interview, phonacoscopy, blood pressure, pulse rate, body temperature, and 12-lead electrocardiogram were performed before and 2 hours after administration. Follow-up visits were scheduled from day 4 up to week 52 to discuss any changes in concomitant medications, to collect vital signs, review adverse events, and obtain blood samples for immunogenicity analyses.

### Outcomes

Trial outcomes were prespecified in the study protocol and did not change following trial commencement. The primary safety end points were the occurrence of solicited local and systemic adverse events that occurred up to 6 days (Day 7) after study drug administration, and any unsolicited adverse events that occurred up to 4 weeks after study drug administration. Information on all adverse events was collected until week 4. Information on serious adverse events, adverse events of special interest, medically-noteworthy or treatment-emergent adverse events as determined by the principal investigator or sub-investigators, and adverse events leading to study discontinuation were collected until the end of study (week 52) or study discontinuation.

The primary immunogenicity end points were neutralising antibody titres against SARS-CoV-2 variants up to 4 weeks after study drug administration. Pseudovirus neutralising antibody titres were calculated as 50% inhibitory dilution (ID_50_), and changes before and after administration of VLPCOV-01 were compared with placebo and BNT162b2. Neutralising activity was assessed against the SARS-CoV-2 spike protein for Wuhan (wild-type), Delta (B.1.617.2), and Omicron (BA.2) variants. Reported secondary immunogenicity assessments were serum IgG titres against SARS-CoV-2 RBD, IgG subclass fraction, and angiotensin-converting enzyme 2 (ACE2)-binding inhibitory activity up to 52 weeks after study drug administration. For quantification of IgG titres in serum, samples were analysed using the SARS-CoV-2 IgG II Quant assay, which detects IgG antibodies to the RBD of the SARS-CoV-2 spike protein, according to the manufacturer’s instructions (Abbott Laboratories). Inhibition of RBD binding to ACE2 was evaluated by V-PLEX SARS-CoV-2 Panel 7 (Meso Scale Diagnostics, LLC). Reported exploratory end points were intracellular cytokine staining of antigen-specific CD4^+^ T-cells. Details of these assays are provided in the Supplementary Appendix.

### Statistical analysis

This report presents interim analyses following data cutoff at day 29, when the study was unblinded. No participants were excluded from the full analysis set (FAS). The safety evaluable set (SES) included all participants who received the study drug after randomisation. Among the SES, safety assessments for solicited adverse events were performed for the participant population where diary solicited adverse event data were available. Safety analyses were presented as numbers and percentages of participants who experienced solicited local and systemic adverse events up to 6 days after study drug administration (day 1 to day 7), and adverse events up to 4 weeks after study drug administration. Two-sided 95% confidence intervals (CI) were calculated using the Clopper-Pearson method. One-sided 95% CIs were used for the upper limit when the proportion was 0, or the lower limit when the proportion was 1. For immunogenicity results, mean titres were calculated by log transformation of source data, and two-sided 95% CIs were based on the t-distribution of the geometric mean. Comparisons between groups were determined by estimated geometric mean titre ratios and two-sided 95% CIs. Results of immunogenicity testing beyond the primary time point (day 29; week 4) excluded results for participants who were infected with SARS-CoV-2 or who received other authorised COVID-19 vaccines during the observation period (appendix p 11).

The trial is registered at the Japan Registry of Clinical Trials under the registration ID jRCT2051210164.

## Results

A total of 92 participants, 46 in each of the two age groups, underwent randomisation between February 16, 2022, and March 17, 2022. Of the 92 participants who received booster injections, 12 received placebo, 20 received 30 μg BNT162b2, 20 received 3 μg VLPCOV-01, 20 received 1 μg VLPCOV-01, and 20 received 0·3 μg VLPCOV-01. All participants who received injections completed study procedures up to 4 weeks post-study drug administration (figure 1). The demographic characteristics of the participants at enrolment are shown in table 1.

**Table 1:**
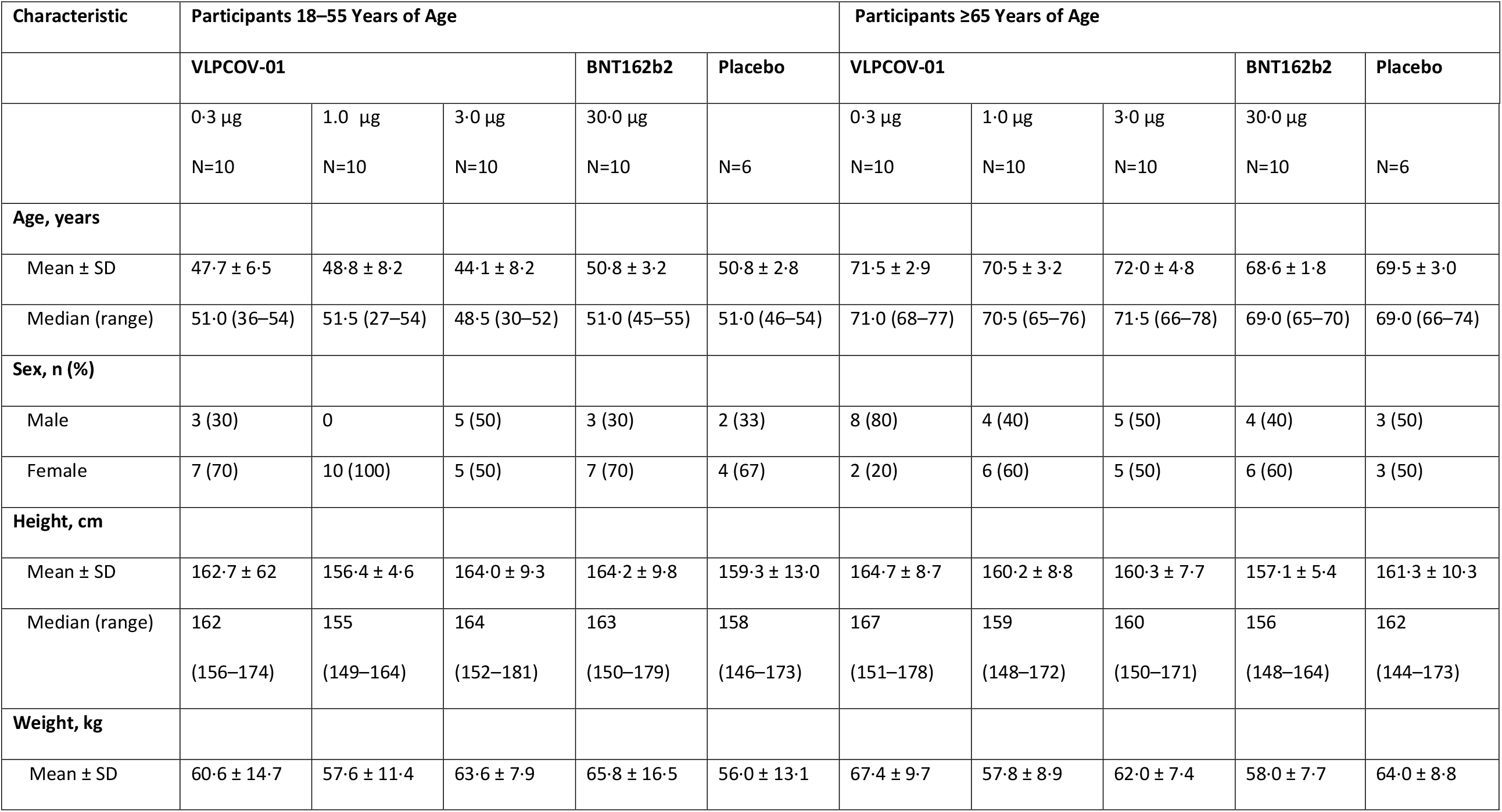
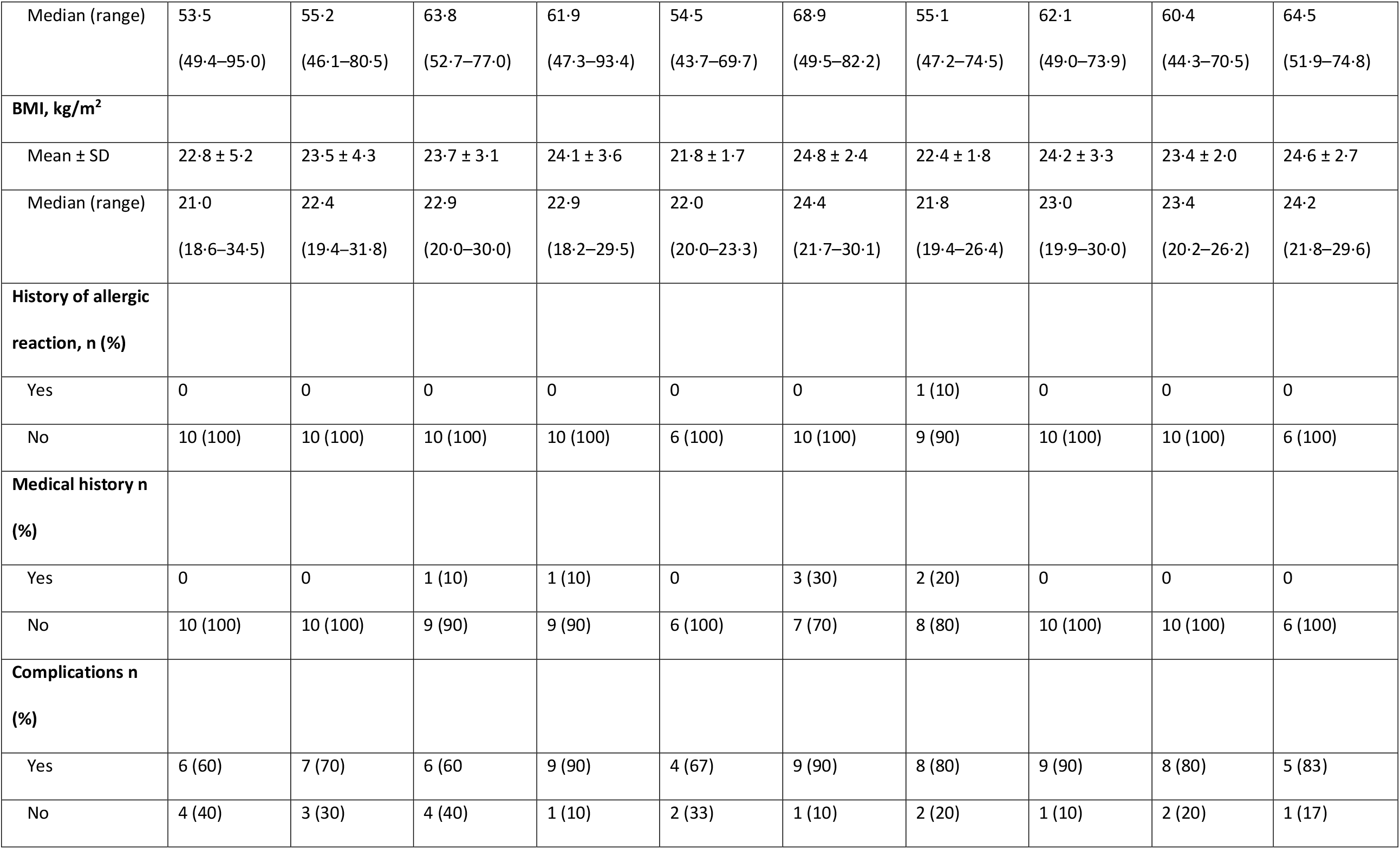
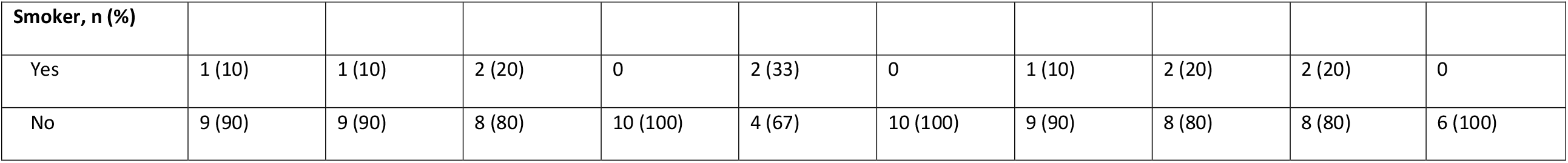
Demographic characteristics of the participants at enrolment.

| Characteristic | Participants 18–55 Years of Age |  |  |  |  | Participants ≥65 Years of Age |  |  |  |  |
| --- | --- | --- | --- | --- | --- | --- | --- | --- | --- | --- |
|  | VLPCOV-01 |  |  | BNT162b2 | Placebo | VLPCOV-01 |  |  | BNT162b2 | Placebo |
|  | 0.3 µg<br>N=10 | 1.0 µg<br>N=10 | 3.0 µg<br>N=10 | 30.0 µg<br>N=10 | N=6 | 0.3 µg<br>N=10 | 1.0 µg<br>N=10 | 3.0 µg<br>N=10 | 30.0 µg<br>N=10 | N=6 |
| **Age, years** |  |  |  |  |  |  |  |  |  |  |
| Mean ± SD | 47.7 ± 6.5 | 48.8 ± 8.2 | 44.1 ± 8.2 | 50.8 ± 3.2 | 50.8 ± 2.8 | 71.5 ± 2.9 | 70.5 ± 3.2 | 72.0 ± 4.8 | 68.6 ± 1.8 | 69.5 ± 3.0 |
| Median (range) | 51.0 (36–54) | 51.5 (27–54) | 48.5 (30–52) | 51.0 (45–55) | 51.0 (46–54) | 71.0 (68–77) | 70.5 (65–76) | 71.5 (66–78) | 69.0 (65–70) | 69.0 (66–74) |
| **Sex, n (%)** |  |  |  |  |  |  |  |  |  |  |
| Male | 3 (30) | 0 | 5 (50) | 3 (30) | 2 (33) | 8 (80) | 4 (40) | 5 (50) | 4 (40) | 3 (50) |
| Female | 7 (70) | 10 (100) | 5 (50) | 7 (70) | 4 (67) | 2 (20) | 6 (60) | 5 (50) | 6 (60) | 3 (50) |
| **Height, cm** |  |  |  |  |  |  |  |  |  |  |
| Mean ± SD | 162.7 ± 6.2 | 156.4 ± 4.6 | 164.0 ± 9.3 | 164.2 ± 9.8 | 159.3 ± 13.0 | 164.7 ± 8.7 | 160.2 ± 8.8 | 160.3 ± 7.7 | 157.1 ± 5.4 | 161.3 ± 10.3 |
| Median (range) | 162<br>(156–174) | 155<br>(149–164) | 164<br>(152–181) | 163<br>(150–179) | 158<br>(146–173) | 167<br>(151–178) | 159<br>(148–172) | 160<br>(150–171) | 156<br>(148–164) | 162<br>(144–173) |
| **Weight, kg** |  |  |  |  |  |  |  |  |  |  |
| Mean ± SD | 60.6 ± 14.7 | 57.6 ± 11.4 | 63.6 ± 7.9 | 65.8 ± 16.5 | 56.0 ± 13.1 | 67.4 ± 9.7 | 57.8 ± 8.9 | 62.0 ± 7.4 | 58.0 ± 7.7 | 64.0 ± 8.8 |
| Median (range) | 53·5<br>(49·4–95·0) | 55·2<br>(46·1–80·5) | 63·8<br>(52·7–77·0) | 61·9<br>(47·3–93·4) | 54·5<br>(43·7–69·7) | 68·9<br>(49·5–82·2) | 55·1<br>(47·2–74·5) | 62·1<br>(49·0–73·9) | 60·4<br>(44·3–70·5) | 64·5<br>(51·9–74·8) |
| **BMI, kg/m<sup>2</sup>** |  |  |  |  |  |  |  |  |  |  |
| Mean ± SD | 22·8 ± 5·2 | 23·5 ± 4·3 | 23·7 ± 3·1 | 24·1 ± 3·6 | 21·8 ± 1·7 | 24·8 ± 2·4 | 22·4 ± 1·8 | 24·2 ± 3·3 | 23·4 ± 2·0 | 24·6 ± 2·7 |
| Median (range) | 21·0<br>(18·6–34·5) | 22·4<br>(19·4–31·8) | 22·9<br>(20·0–30·0) | 22·9<br>(18·2–29·5) | 22·0<br>(20·0–23·3) | 24·4<br>(21·7–30·1) | 21·8<br>(19·4–26·4) | 23·0<br>(19·9–30·0) | 23·4<br>(20·2–26·2) | 24·2<br>(21·8–29·6) |
| **History of allergic reaction, n (%)** |  |  |  |  |  |  |  |  |  |  |
| Yes | 0 | 0 | 0 | 0 | 0 | 0 | 1 (10) | 0 | 0 | 0 |
| No | 10 (100) | 10 (100) | 10 (100) | 10 (100) | 6 (100) | 10 (100) | 9 (90) | 10 (100) | 10 (100) | 6 (100) |
| **Medical history n (%)** |  |  |  |  |  |  |  |  |  |  |
| Yes | 0 | 0 | 1 (10) | 1 (10) | 0 | 3 (30) | 2 (20) | 0 | 0 | 0 |
| No | 10 (100) | 10 (100) | 9 (90) | 9 (90) | 6 (100) | 7 (70) | 8 (80) | 10 (100) | 10 (100) | 6 (100) |
| **Complications n (%)** |  |  |  |  |  |  |  |  |  |  |
| Yes | 6 (60) | 7 (70) | 6 (60) | 9 (90) | 4 (67) | 9 (90) | 8 (80) | 9 (90) | 8 (80) | 5 (83) |
| No | 4 (40) | 3 (30) | 4 (40) | 1 (10) | 2 (33) | 1 (10) | 2 (20) | 1 (10) | 2 (20) | 1 (17) |

| Smoker, n (%) |  |  |  |  |  |  |  |  |  |  |
| --- | --- | --- | --- | --- | --- | --- | --- | --- | --- | --- |
| Yes | 1 (10) | 1 (10) | 2 (20) | 0 | 2 (33) | 0 | 1 (10) | 2 (20) | 2 (20) | 0 |
| No | 9 (90) | 9 (90) | 8 (80) | 10 (100) | 4 (67) | 10 (100) | 9 (90) | 8 (80) | 8 (80) | 6 (100) |

**Figure 1:**
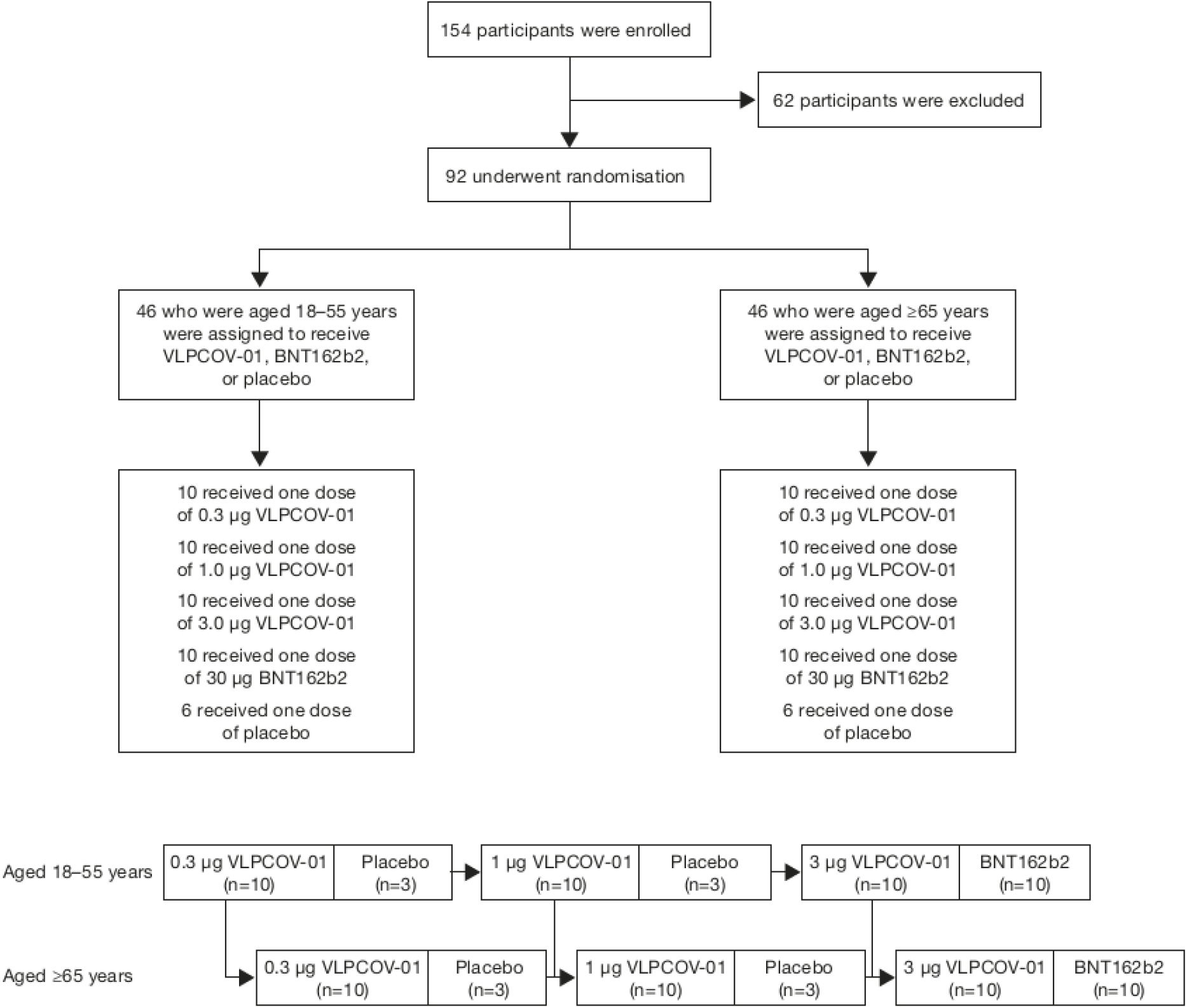
Participant randomisation and schedule. The full analysis set and safety analysis set included all participants who underwent randomisation and who received one dose of VLPCOV-01, BNT162b2, or placebo. All participants who received injections completed study procedures up to 4 weeks post-study drug administration. For VLPCOV-01 administration, transition from the low dose cohort to the next dose cohort in the same age group, and from the non-elderly cohort to the elderly cohort, was performed after the principal investigator determined that there were no medical concerns up to day 4.

No serious adverse events were reported, and no prespecified trial-halting events were met. Following booster vaccination with VLPCOV-01, the majority of solicited adverse events reported were mild to moderate in severity and generally comparable to booster vaccination with BNT162b2 (figure 2). Among participants in the non-elderly cohort, two participants (20%) who received 3 μg VLPCOV-01 and two participants (20%) who received 30 μg BNT162b2 experienced solicited local adverse events (pain [3 μg VLPCOV-01] and tenderness [30 μg BNT162b2]) that were rated as severe. Two participants (20%) who received 3 μg VLPCOV-01, two participants (20%) who received 1 μg VLPCOV-01, and one participant (10%) who received 30 μg BNT162b2 experienced solicited systemic adverse events (fatigue [3 μg VLPCOV-01], myalgia [3 μg and 1 μg VLPCOV-01], chills [3 μg and 1 μg VLPCOV-01 and 30 μg BNT16b2], and arthralgia [3 μg and 1 μg VLPCOV-01]) that were rated as severe. No solicited adverse events reported by participants aged ≥65 years in the study were classified as severe (figure 2).

**Figure 2:**
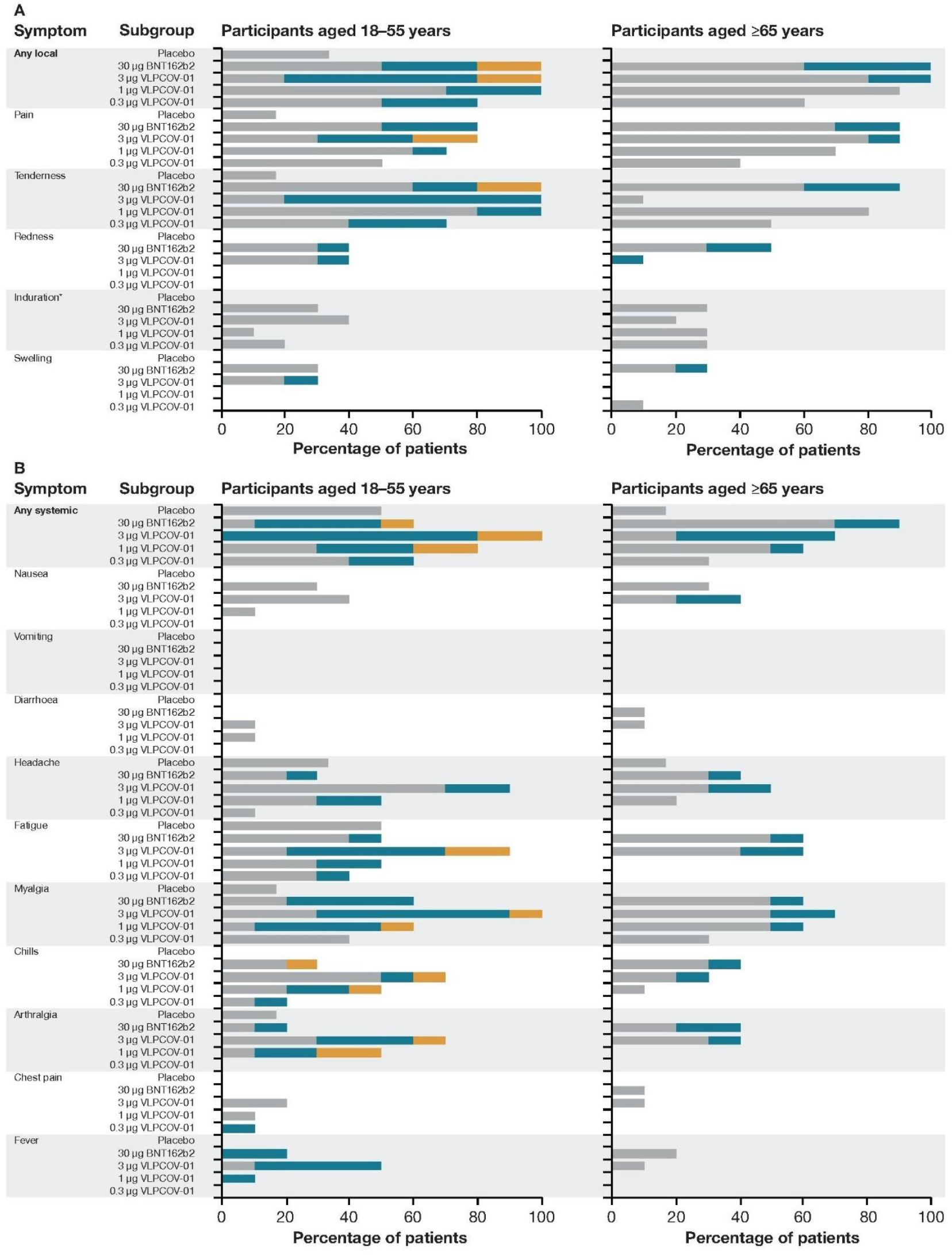
Solicited adverse events reported up to 6 days after study drug administration (day 7) Panel A shows the percentage of solicited local adverse events and their severity reported up to 6 days after study drug administration. Panel B shows the percentage of systemic adverse events and their severity reported up to 6 days after study drug administration. The severity of solicited adverse events was graded as mild, moderate, or severe based on criteria that are described in the appendix p 9–10. *No severity assessment was made for induration, only a yes or no assessment was made. Percentages shown are participants that recorded “yes”.

The most common solicited adverse events experienced following VLPCOV-01 vaccination among participants aged 18 to 55 years were pain, tenderness, myalgia, fatigue, headache, chills, and arthralgia, and among participants aged ≥65 years were pain, tenderness, and myalgia (figure 2). Local and systemic reactogenicity events following vaccination with VLPCOV-01 typically occurred on the day of vaccination or one day afterward and resolved by day 7 (appendix p 12–20).

Unsolicited adverse events and clinical laboratory values revealed no patterns of concern and are detailed in appendix p 21–22.

Among non-elderly participants, VLPCOV-01 induced IgG titres against SARS-CoV-2 RBD protein (Wuhan variant) that were maintained up to 26 weeks post-vaccination with all doses (figure 3A). In the non-elderly cohort, IgG titres at 13 weeks post-vaccination in the 3 μg VLPCOV-01 group were 20833 (95% CI 8467 to 51261) compared to 6924·34 (95% CI 3988 to 12021) in the 30 μg BNT162b2 group. IgG titres remained high at 26 weeks post-vaccination with 3 μg VLPCOV-01, which continued to be higher than IgG titres measured following vaccination with 30 μg BNT162b2 (12873 [95% CI 937 to 17686] vs. 3166 [95% CI 1619 to 6191]). IgG titres at 26 weeks post-vaccination with 1 μg (9282 [95% CI 2753 to 31299]) and 0·3 μg (5037 [95% CI 1272 to 19940]) VLPCOV-01 were also higher than IgG titres following vaccination with 30 μg BNT162b2.

**Figure 3:**
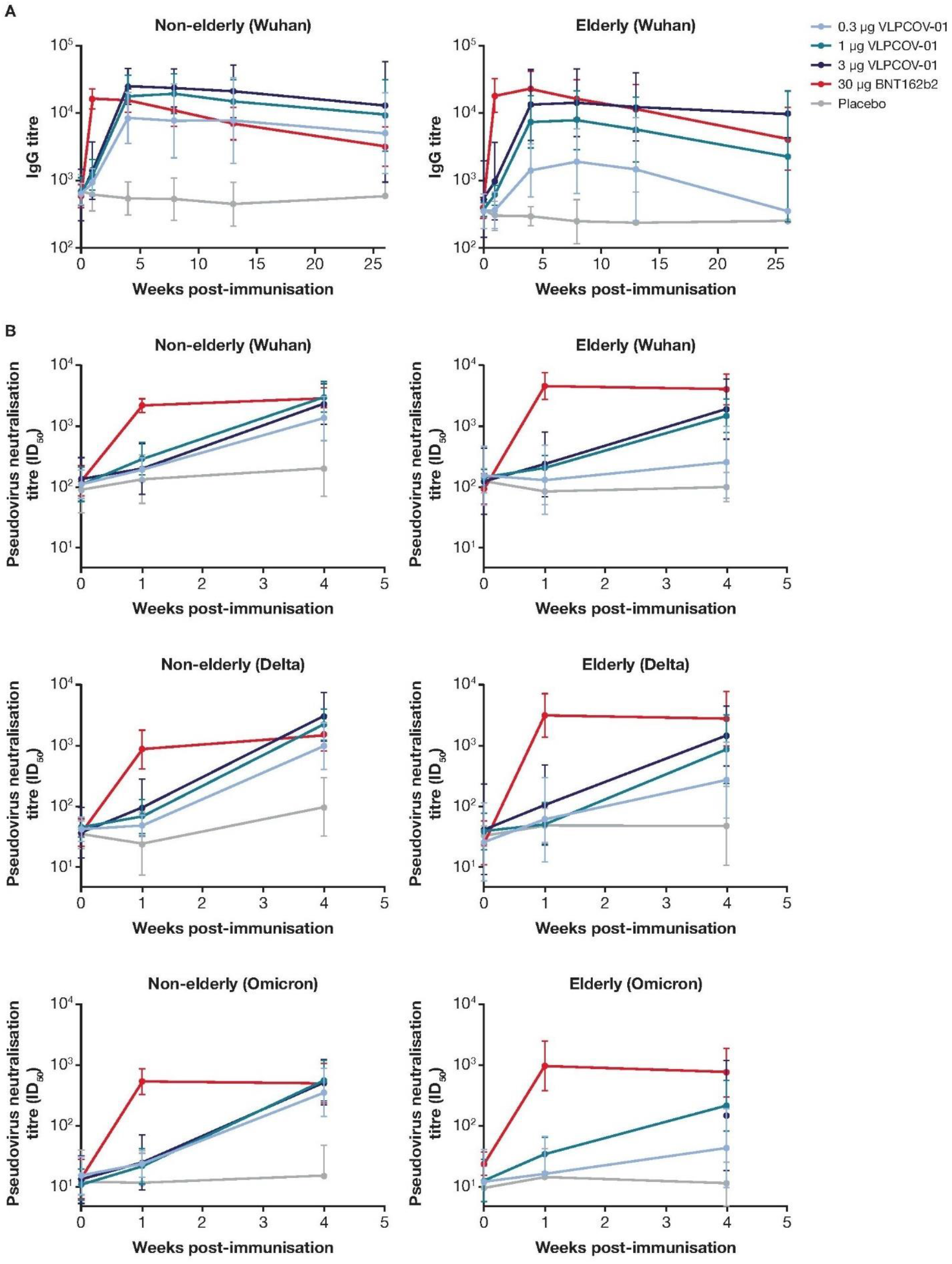
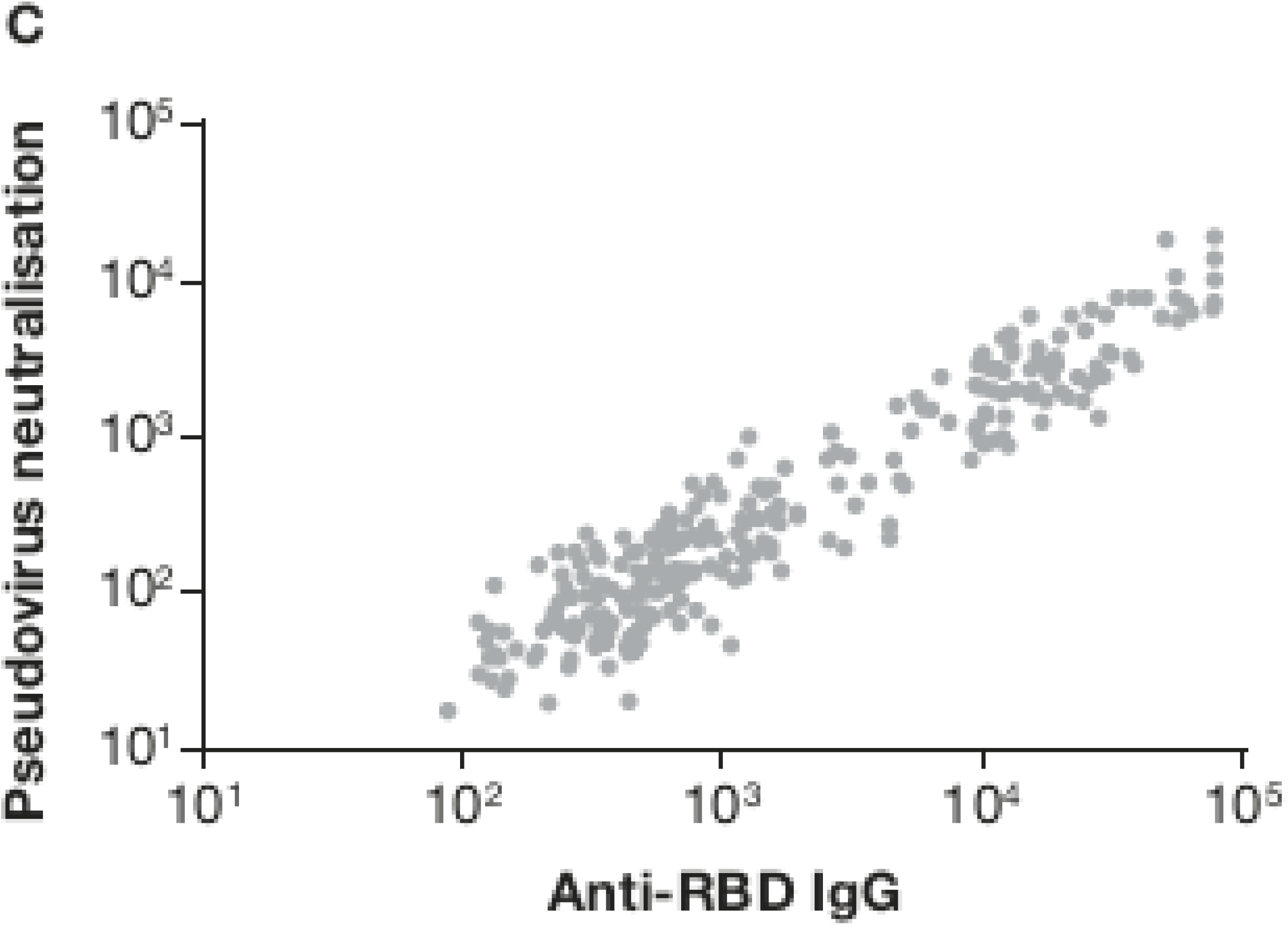
SARS-CoV-2 IgG and neutralising antibody responses. Panel A shows serum IgG titres against wild type SARS-CoV-2 RBD protein and Panel B shows pseudovirus neutralising antibody titres (ID_50_) against SARS-CoV-2 variants for non-elderly (left side) and elderly (right side) participants. Participants received one injection of VLPCOV-01 (0·3, 1·0, or 3·0 μg), 30 μg BNT162b2, or placebo on day 1 (week 0). Logarithmic values are reported as geometric mean titres for serum IgG and neutralising antibody against pseudovirus. Bars indicate 95% CIs. Panel C shows the correlation between serum neutralising antibody titres against pseudovirus (wild type) and IgG antibody titres against SARS-CoV-2 RBD protein following booster vaccination. Pearson’s productmoment correlation coefficient and p value were calculated following log transformation of source data (*r*=0·950, p<0·001).

In the elderly cohort, all doses of VLPCOV-01 induced IgG titres that were maintained up to 13 weeks post-vaccination. IgG titres at 26 weeks post-vaccination with 3 μg VLPCOV-01 were 9865 (95% CI 4396 to 22138) compared to 4183 (95% CI 1436 to 12180) following vaccination with 30 μg BNT162b2.

SARS-CoV-2 neutralisation titres were comparable for all groups at baseline. Neutralising antibody titres against all variants of SARS-CoV-2 tested were induced by all doses of VLPCOV-01 in participants from both age cohorts, and a dose effect was seen (figure 3B). At 4 weeks post-vaccination (day 29), results were comparable between 1 μg and 3 μg VLPCOV-01 and 30 μg BNT162b2. Additionally, inhibition of spike-ACE2 binding and RBD-ACE2 binding for all variants was observed at 4 weeks post-vaccination with all concentrations of VLPCOV-01, and in both age cohorts (figure S1).

A strong correlation was observed between anti-RBD IgG titres and pseudovirus-neutralising responses to VLPCOV-01 booster vaccination (figure 3C; *r*=0·950, p<0·001).

In response to RBD-specific peptide pools, CD4^+^ T-cell responses were induced by booster vaccination with VLPCOV-01 among participants aged 18 to 55 years and ≥65 years (figure 4). Across all doses of VLPCOV-01, responses were Th1 dominant (figure 4B), with minimal Th2 (figure 4C) and Th17 (figure 4D) responses. Responses to spike-specific peptide pools (figure S2A) revealed that, in comparison to 3 μg VLPCOV-01, 30 μg BNT162b2 induced stronger spike-specific Th1 responses (among non-elderly, geometric mean 1·18 vs. 1·98 fold-change; among elderly, geometric mean 1·27 vs. 1·45 fold-change), Th2 responses (among non-elderly, geometric mean 1·18 vs. 2·43 fold-change; among elderly, geometric mean 1·20 vs. 1·60 fold-change), and IL-21 responses (among non-elderly, geometric mean 1·49 vs. 8·14 fold-change; among elderly, geometric mean 1·70 vs. 4·96 fold-change). Absolute percentages of RBD-specific CD4^+^ T-cell responses are shown in appendix p 7.

**Figure 4:**
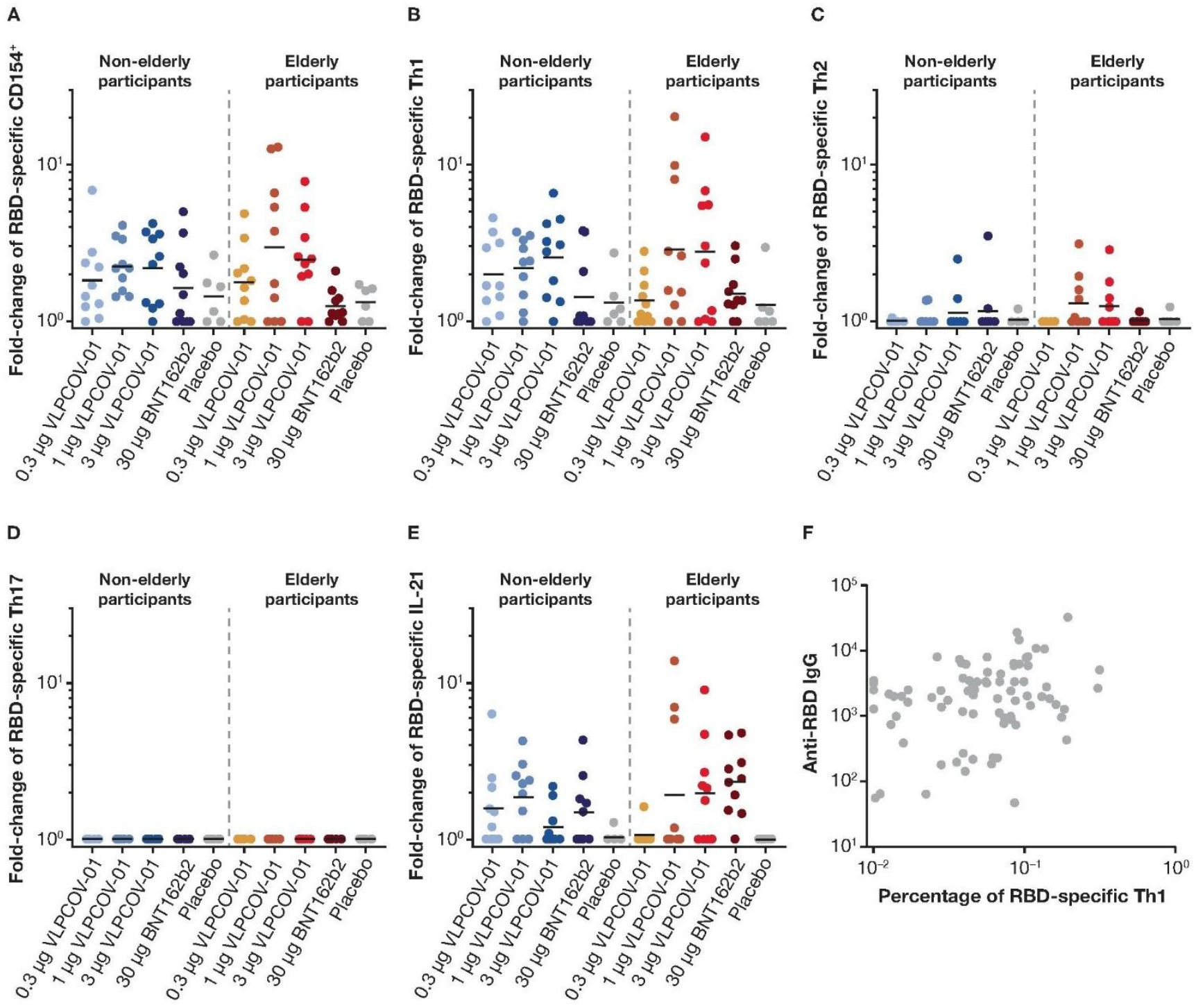
CD4^+^ T-cell responses. Flow cytometric analysis was performed to analyse RBD-specific T cells. Responses to VLPCOV-01, BNT162b2, or placebo are shown as fold-change from baseline (day 1) to week 4 (day 29) for each cohort. Panel B shows the response in CD4^+^ Th1 cells was characterised by the expression of interleukin-2, tumour necrosis factor α, and interferon-γ. Panel C shows the response in Th2 cells, which was measured by expression of interleukin-4 and interleukin-13. For Panels A–E, the horizontal bars indicate median values. Panel F shows the correlation between IgG antibody titres against SARS-CoV-2 RBD protein and the percentage of RBD-specific Th1 cell responses following vaccination with VLPCOV-01. Spearman’s rank correlation coefficient and p value were calculated following log transformation of source data (*r*=0·2717, p=0·0092).

Exploratory analysis demonstrated correlation between RBD-specific Th1 responses and neutralising antibody titres 4 weeks after booster vaccination with VLPCOV-01 (figure 4F; *r=*0·2717, p=0·0092). Furthermore, analyses of the distribution of IgG subclasses induced by VLPCOV-01 vaccination identified dominant IgG1 responses that were comparable to those induced by BNT162b2 at 4 weeks post-vaccination (figure S3).

## Discussion

Here, we present the findings from the first clinical trial of a saRNA vaccine expressing an RBD-anchored antigen. The primary safety and immunogenicity analyses from this phase 1 clinical trial of VLPCOV-01 indicate that in healthy adult participants ≥18 years who have previously received primary COVID-19 vaccination, VLPCOV-01 had an acceptable safety profile and induced high immune responses in both non-elderly and elderly participants, with neutralising antibody activity strongly correlating with anti-SARS-CoV-2 IgG.

Since the first reported cases of COVID-19 in December 2019, multiple SARS-CoV-2 variants with increased transmissibility and greater antibody escape have emerged.^2^ The Omicron variant and its sublineages are highly transmissible, with multiple mutations in the spike protein that result in escape from neutralising antibody responses that are elicited by COVID-19 vaccination.^15,16^ Booster vaccination may lead to an increase in neutralising antibody titres against Omicron, however, it has been shown that these responses wane after a third dose of mRNA vaccine.^16,17^ Mutations in SARS-CoV-2 variants that occur in the spike N-terminal domain or in the RBD, regions that are critical for ACE2 binding, may compromise neutralising antibody activity.^18^ In this study, we demonstrated that VLPCOV-01 booster vaccination elicited broad neutralising antibody responses, with neutralising antibody titres to wild-type, Delta, and Omicron variants observed. Furthermore, the neutralising antibodies were capable of blocking the spike-ACE2 and RBD-ACE2 interactions for all SARS-CoV-2 variants tested.

The COVID-19 mRNA vaccines BNT162b2 and mRNA-1273 have demonstrated waning immunity, with high initial neutralising antibody titres declining by 3 to 6 months, necessitating the need for booster vaccinations.^19-21^ In this interim analysis, we describe neutralising activity up to week 4. Further long-term analysis is required to ascertain the duration of response induced by VLPCOV-01, however, the strong correlation of anti-SARS-CoV-2 IgG titres with neutralisation suggests that IgG titres may be used as a predictor of neutralising activity. High IgG titres were maintained up to 13 weeks post-vaccination with VLPCOV-01, and IgG titres measured from samples collected 26 weeks post-vaccination with 3 μg VLPCOV-01 suggest that these may be maintained beyond 6 months.

In addition to robust humoral responses, we also demonstrated the ability of VLPCOV-01 to induce RBD-specific CD4^+^ T-cell responses that were comparable to BNT162b2. Although whole spike-specific CD4^+^ T-cell responses were higher in BNT162b2 compared to VLPCOV-01 (figure S2B), correlation between RBD-specific Th1 responses and neutralising antibody titres 4 weeks following booster vaccination with VLPCOV-01 (figure 4) suggest that CD4^+^ T-cell induction is sufficient to help maintain neutralising activity. Moreover, total RBD-specific CD4^+^ T-cells (CD154^+^) were dominated by Th1 cells, which is consistent with the reported T-cell responses for other COVID-19 vaccines.^22-26^

Studies have demonstrated the importance of vaccine-elicited T-cell responses in coordinating humoral and cellular immune responses and their extended durability despite declining antibody titres.^27,28^ This suggests that T-cell responses may be critical for continued protection against severe disease in the absence of neutralising antibodies. The VLPCOV-01-induced T-cell responses that formed across both age cohorts may therefore provide critical protection from severe illness due to SARS-CoV-2 infection.

Limitations of this study include the relatively short follow-up to date and small trial size, particularly at later immunogenicity analyses time points, where the number of participants in the analysis decreased over time due to vaccination with authorised vaccines post-week 4 or infection with SARS-CoV-2.Adverse event reporting is ongoing and will continue until the planned study duration of 52 weeks. We are conducting a phase 2 dose-ranging study to further evaluate the immunogenicity response and safety profile of VLPCOV-01.

There are other COVID-19 vaccines in clinical development that either utilise the saRNA vaccine platform^11^ or specifically target the RBD of the spike protein.^29^ However, to our knowledge, this is the first report from a clinical study of a COVID-19 vaccine that encompasses both the saRNA platform and RBD-anchored antigen design. We have shown that VLPCOV-01 has a favourable safety profile and induces immune responses that are comparable to the BNT162b2 mRNA vaccine. Importantly, the responses induced by VLPCOV-01 are achieved at 1/10 of the BNT162b2 dose with potentially longer duration, which is advantageous when considering the importance of being able to produce vaccines in large quantities to meet the demands of the COVID-19 pandemic. A low-dose vaccine such as VLPCOV-01 enables production of more doses for the same amount of vaccine, improving manufacturing scale and time and increasing distribution volume. Overall, the results from this study suggest that VLPCOV-01 could be used as an alternative to the mRNA vaccines that are currently available for COVID-19 booster vaccination, and support further development of this vaccine.

## Data Availability

All data produced in the present work are contained in the manuscript

## Contributors

T Sekida, N sato, Y Iwatani, T Fukuhara, T Yamamoto, J F Smith, and W Akahata conceived, designed, and coordinated the study. T Nogimori, A Washizaki, and Y Masuta performed T-cell analysis. H Ode, T Tamura, and R Suzuki performed IgG analysis. Y Kazami, N Sato, K Kono, T Sekida, M Nakata and T Hasunuma prepared and executed the clinical study. M Komori, K Ishimoto, A Morey, K Matsuda and W Akahata contributed to the data validation, analysis, writing and revision of the manuscript. All authors read and approved the manuscript, had full access to all the data in the study and had final responsibility for the decision to submit for publication.

## Data sharing

Access to related study documents (e.g., study protocol, statistical analysis plan, clinical study report) will be provided upon request from qualified researchers, and subject to certain criteria, conditions, and exceptions.

## Acknowledgments

This study was supported by AMED under Grant Number JP21nf0101627. Medical writing support was provided by Emily Feist PhD of Parexel International, which was funded by VLP Therapeutics Japan, LLC.

## Supplemental Appendix

### Supplemental methods

#### Assessment of T-cell responses

For analyzing antigen-specific T cells, flow cytometric analysis was performed. The cell staining protocol has been previously described.^1^ Briefly, peripheral blood mononuclear cells (PBMCs) obtained from participants were incubated in 200 μl RPMI medium containing 10% FBS with or without peptides (17-mers overlapping by 10 residues) corresponding to the RBD region or the full SARS-CoV-2 spike region, at a final concentration of 2 μg/ml of each peptide in the presence of anti-CD107a (H4A3). Thereafter, 0.2 μl BD GolgiPlug and 0.14 μl BD GolgiStop (both from BD Biosciences) were added to the cells and the cells were incubated for 5.5 h. The cells were then stained using the LIVE/DEAD™ Fixable Blue Dead Cell Stain Kit (Thermo Fisher Scientific) and stained with anti-CD3 (SP34-2), anti-CD4 (S3.5), anti-CD8 (RPA-T8), anti-CD27 (1A4CD27), and anti-CD45RO (UCHL1) antibodies. After fixation and permeabilization using the Cytofix/Cytoperm kit (BD Biosciences), the cells were stained with anti-IFN-g (4S.B3), anti-TNF (MAb11), anti-CD154 (TRAP1), anti-IL-13 (JES10-5A2), anti-IL-21 (3A3-N21), anti-IL-4 (8D4-8), anti-IL-17A (BL168), and anti-IL-2 (MQ1-17H12) antibodies. After washing, the cells were fixed with 1% paraformaldehyde and analyzed using a FACSymphony A5 instrument equipped with five lasers (BD Biosciences). Data were analyzed using the FlowJo software version 10.7.1 (BD Biosciences).

#### Pseudovirus production

The DNA sequences of SARS-CoV-2 spike gene (Wuhan spike: NC_045512.2, Delta spike: QWA53965.1, Omicron BA.2 spike: UFO69279.1) were codon-optimized for human cells and inserted into eukaryotic expression vactor pCAGG to generate the envelope plasmids. The envelop plasmid was transfected into HEK293T cells by TransIT-LT1 (Mirus Bio) and incubated for 24h at 37 °C. The cells were infected with the VSVΔG-Luc/G, in which the G envelope was replaced with the reporter luciferase gene, and which was pseudo-typed with the VSV-G glycoprotein.^2,3^ The virus was absorbed, washed, and incubated for 24 h at 37 °C. The culture supernatant was collected and stored at −80 °C after removal of cell debris by centrifugation.

#### Pseudovirus neutralization assay

The Vero cells (1.5 × 10^4^ cells per well) were seeded on 96-well plate and incubated overnight at 37 °C. The serum samples were inactivated at 56 °C for 30 min and diluted from 10 to 40,960 dilution with DMEM medium. 60 μL of diluted serum sample was mixed with the equal volume of pseudo-typed virus (equivalent to 2.5 × 10^6^ RLU/mL) for 1h at 37 °C. Then, 100 μL of mixture (serum/pseudo virus) was added to the Vero cells and incubated for 24 h at 37 °C. The cells were lysed and activated with the Luciferase Assay system (Promega). Luciferase activity of the cells were measured by Synergy LC (Bio Tek). The neutralization activity was analyzed by using GraphPad Prism 8. RLU reduction (percentage) was calculated as: 1-(RLU of samples – RLU of pseudo-typed virus only wells)/(RLU from medium only wells) × 100 (%). The neutralization titer was calculated as 50 % inhibitory dilution (ID_50_).

#### ACE2 binding inhibition

Inhibition of ACE2 binding to SARS-CoV-2 S1 RBD (wild-type, B.1.1.7 and P.1), and SARS-CoV-2 Spike (wild-type, B.1.1.7, B.1.351 and P.1) were measured using the V-PLEX COVID-19 Coronavirus Panel 7 (ACE2) multiplexed immunoassay kit (Meso Scale Diagnostics, Rockville, MD USA. Cat.# K15440U-2). The assays were performed according to manufacturer’s instructions. Briefly, antigen-coated 96-well plates were blocked with MSD Blocker A for 30 minutes. Following 3 washes with MSD wash buffer, samples (diluted 1:10 or 1:80 in diluent buffer) or standard solutions were added to the wells. After 2-h incubation, ACE2 Detection Solution (SULFO-TAG Human ACE2 Protein, 1/200 dilution, Cat.# D21ADG-3) was added. Following 3 washes, MSD GOLD Read Buffer B was added to the wells, and plates were read using a MESO QuickPlex SQ 120MM Reader. The standard curve was established by fitting the signals from the standard using a 4-parameter logistic model. Concentrations (Units/mL) of samples were determined from the electrochemiluminescence signals and multiplied by the dilution factor.

**Figure S1.**
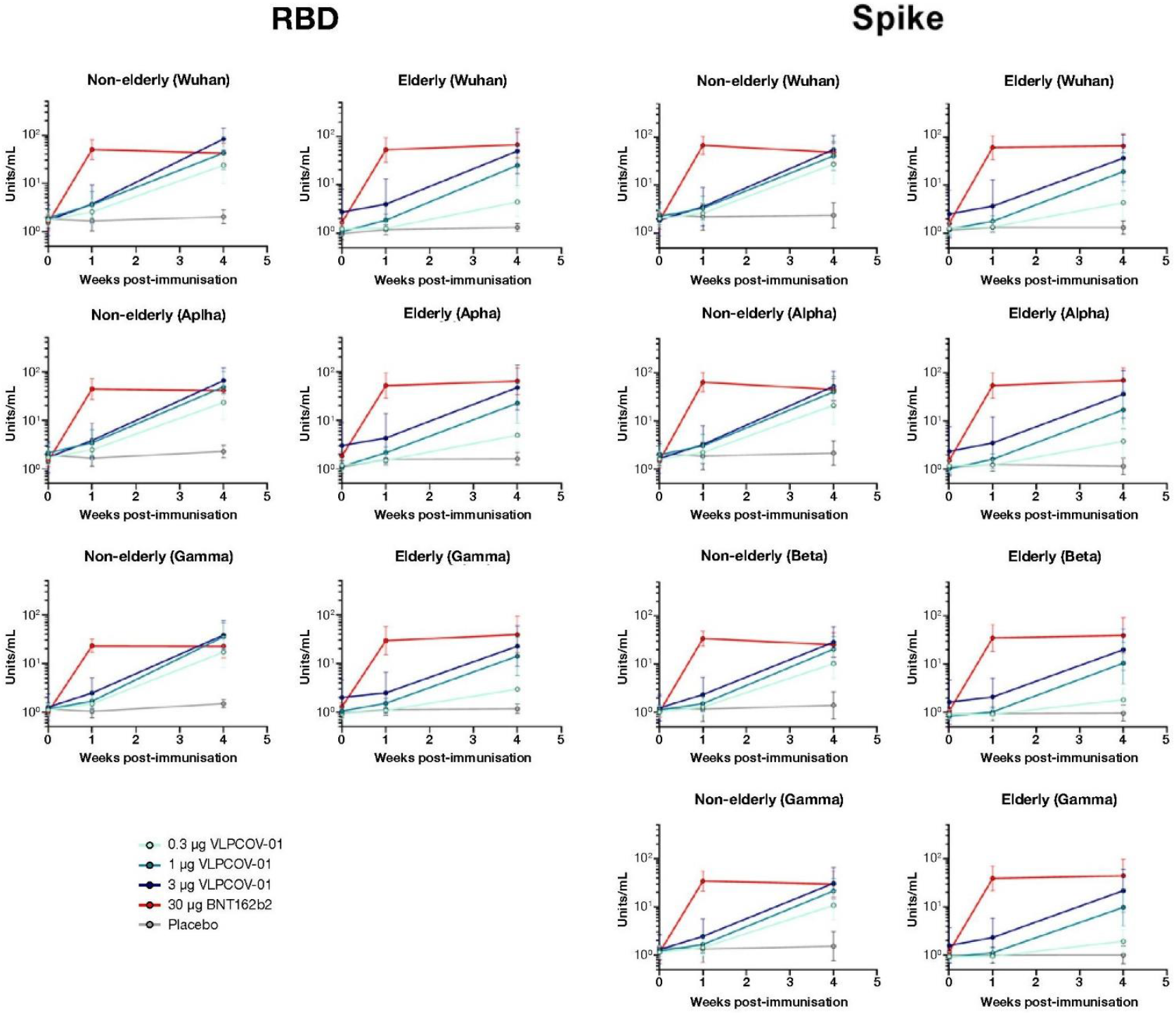
ACE2 Binding Inhibition.

**Figure S2A.**
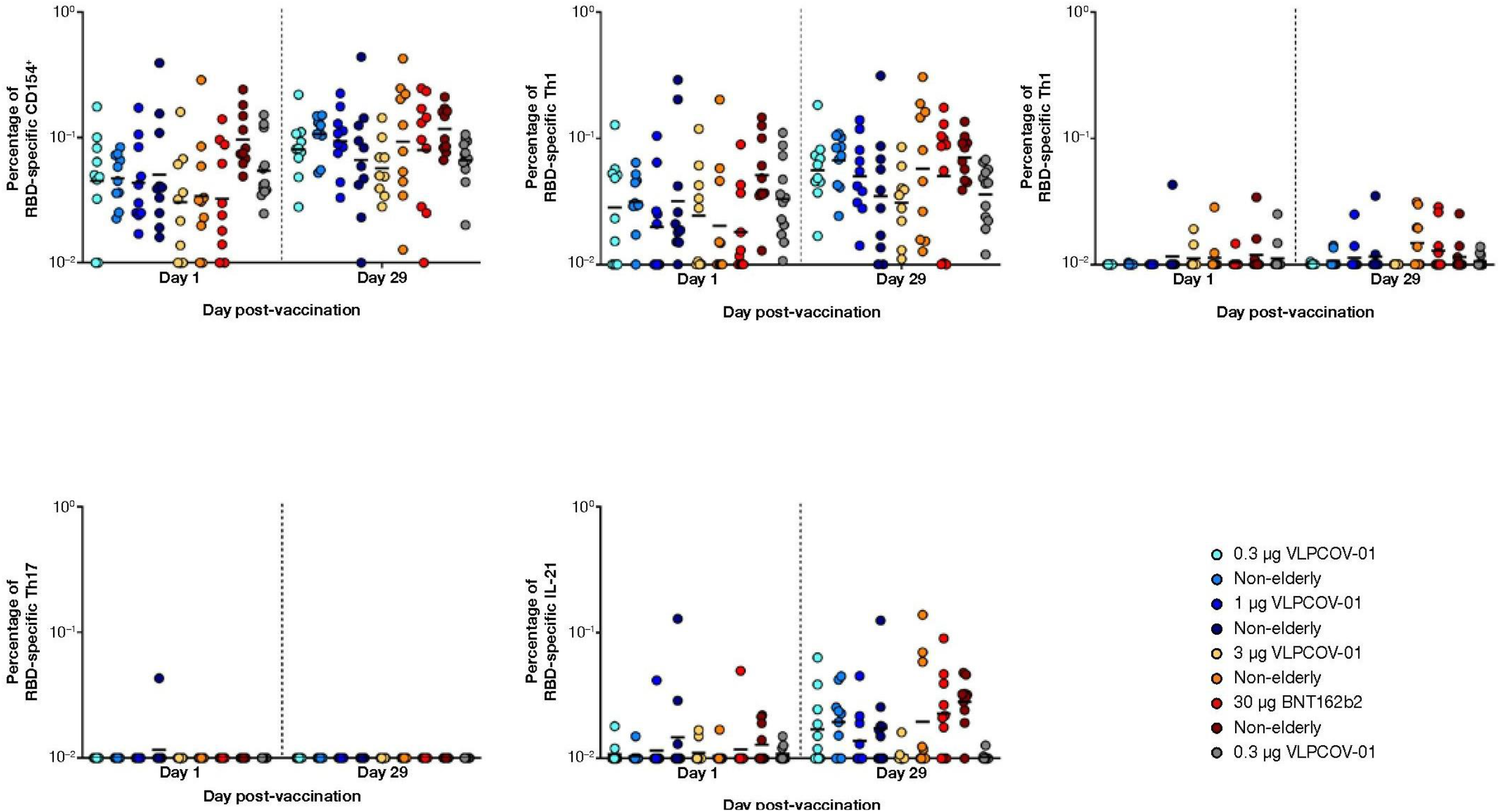
Absolute RBD-specific CD4^+^ T-cell Reponses.

**Figure S2B.**
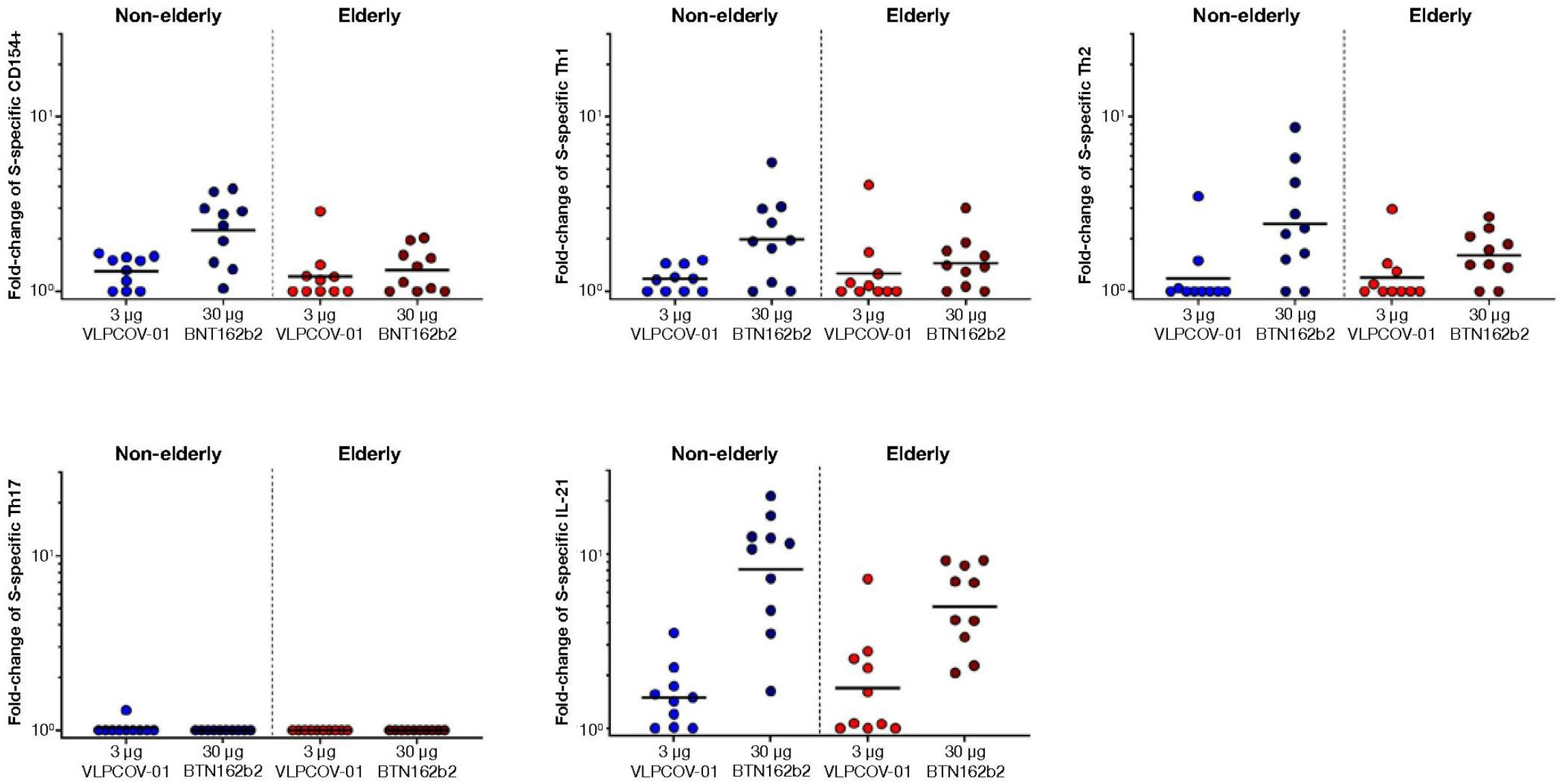
Spike-specific CD4^+^ T-cell Responses.

**Figure S3.**
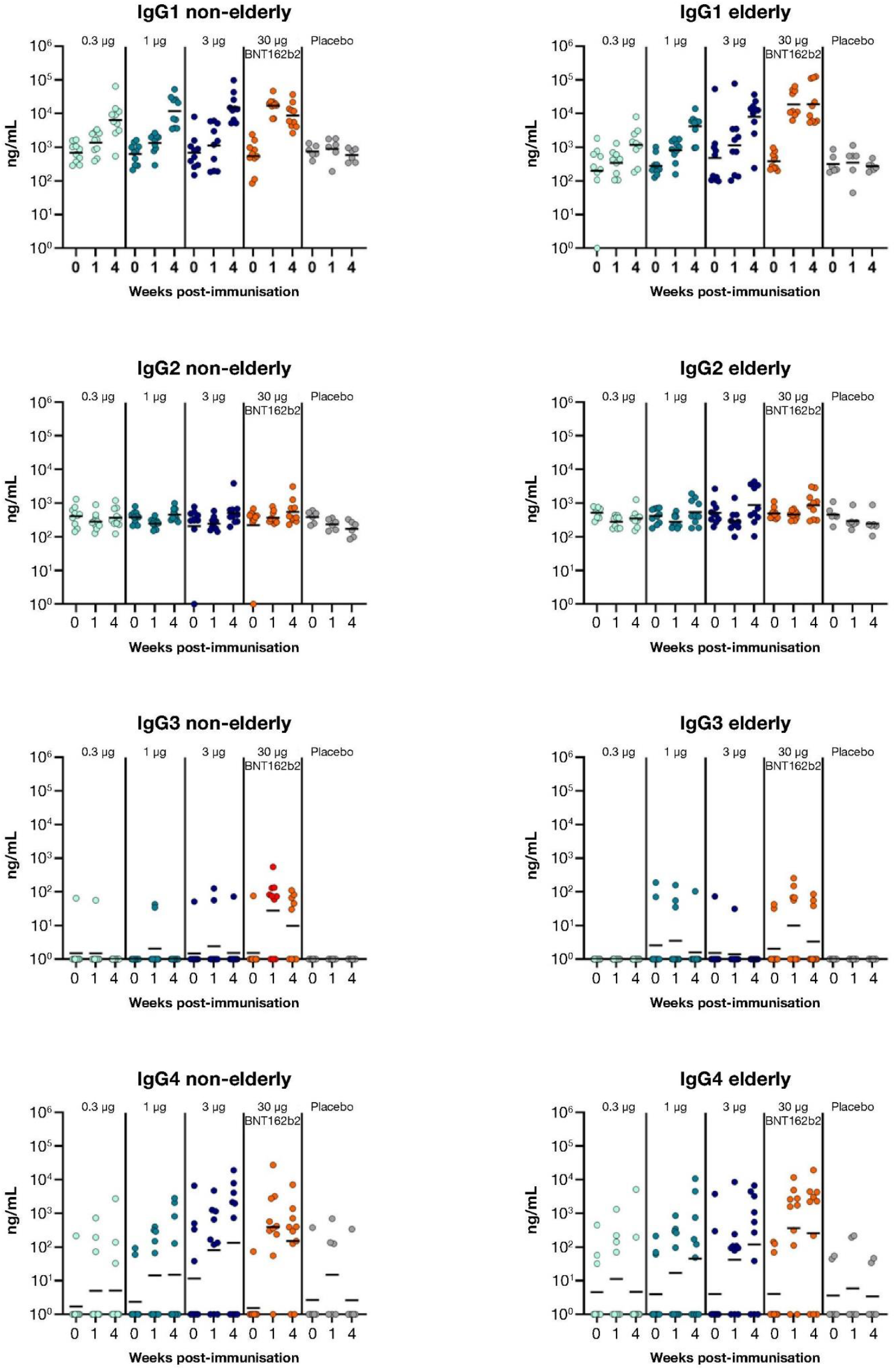
IgG Subclass Analysis.

**Table S1.** Severity of Solicited Adverse Events.

|  |  | Present |  |  |  |
| --- | --- | --- | --- | --- | --- |
|  |  | Mild<br>Grade 1 | Moderate<br>Grade 2 | Severe<br>Grade 3 | Potentially life-threatening<br>Grade 4 |
| Local | Pain | Does not interfere with activities | Interferes with activities | Interferes with daily activities | Emergency transportation or hospitalization due to severe pain |
|  | Tenderness | Mild discomfort when touching | Discomfort when moving | Marked discomfort at rest | Emergency transportation or hospitalization |
|  | Redness | 2.5–5 cm | 5.1–10 cm | >10 cm | Necrosis or exfoliative dermatitis |
|  | Induration | / | / | / | Necrosis |
|  | Swelling | 2.5–5 cm | 5.1–10 cm | >10 cm | Necrosis |
| Systemic | Nausea | Can eat and drink | Some difficulty when eating or drinking | Cannot eat or drink | Emergency transportation or hospitalization due to severe nausea |
|  | Vomiting | 1–2 times in 24 hours | >2 times in 24 hours | IV hydration | Emergency transportation or hospitalization due to hypotensive shock |
|  | Diarrhea | 2-3 loose stools in 24 hours | 4-5 loose stools in 24 hours | ≥6 loose stools in 24 hours | Emergency transportation or hospitalization due to severe diarrhea |
|  | Headache | Does not interfere with activities | Interferes with some activities | Interferes with daily activities | Emergency transportation or hospitalization due to severe headache |
|  | Fatigue | Does not interfere with activities | Interferes with some activities | Interferes with daily activities | Emergency transportation or hospitalization due to severe fatigue |
|  | Myalgia | Does not interfere with activities | Interferes with some activities | Interferes with daily activities | Emergency transportation or hospitalization due to severe new or worsening myalgia |
|  | Chills | Does not interfere with activities | Interferes with some activities | Interferes with daily activities | Emergency transportation or hospitalization due to severe chills |
|  | Arthralgia | Does not interfere with activities | Interferes with some activities | Interferes with daily activities | Emergency transportation or hospitalization due to severe new or worsening arthralgia |
|  | Chest pain | Does not interfere with activities | Interferes with some activities | Interferes with daily activities | Emergency transportation or hospitalization due to severe new or worsening chest pain |
|  | Fever | 38.0–38.4°C | 38.5–38.9°C | 39.0–40°C | >40°C |

**Table S2.** Participant Exclusion During Study Observation Period.

| Participants | Number of participants with<br>SARS-CoV-2 infection<br>(week excluded) | Number of participants who received other<br>authorized COVID-19 vaccines<br>(week excluded) |
| --- | --- | --- |
| **18–55 Years of Age** |  |  |
| 0.3-µg VLPCOV-01 | 1 (week 26) | 3 (2 week 8; 1 week 13) |
| 1-µg VLPCOV-01 | 2 (week 26) | 4 (1 week 8; 3 week 26) |
| 3-µg VLPCOV-01 | 5 (2 week 13; 3 week 26) | 1 (week 26) |
| 30-µg BNT162b2 | 2 (week 26) | 1 (week 8) |
| Placebo | 1 (week 26) | 3 (1 week 8; 2 week 26) |
| **≥65 Years of Age** |  |  |
| 0.3-µg VLPCOV-01 | 2 (1 week 0; 1 week 26) | 7 (2 week 8; 2 week 13; 3 week 26) |
| 1-µg VLPCOV-01 | 2 (week 26) | 3 (1 week 8; 1 week 13; 1 week 26) |
| 3-µg VLPCOV-01 | 1 (week 26) | 1 (week 26) |
| 30-µg BNT162b2 | 1 (week 26) | 3 (1 week 8; 2 week 26) |
| Placebo | 0 | 5 (3 week 8; 2 week 26) |

**Table S3.**
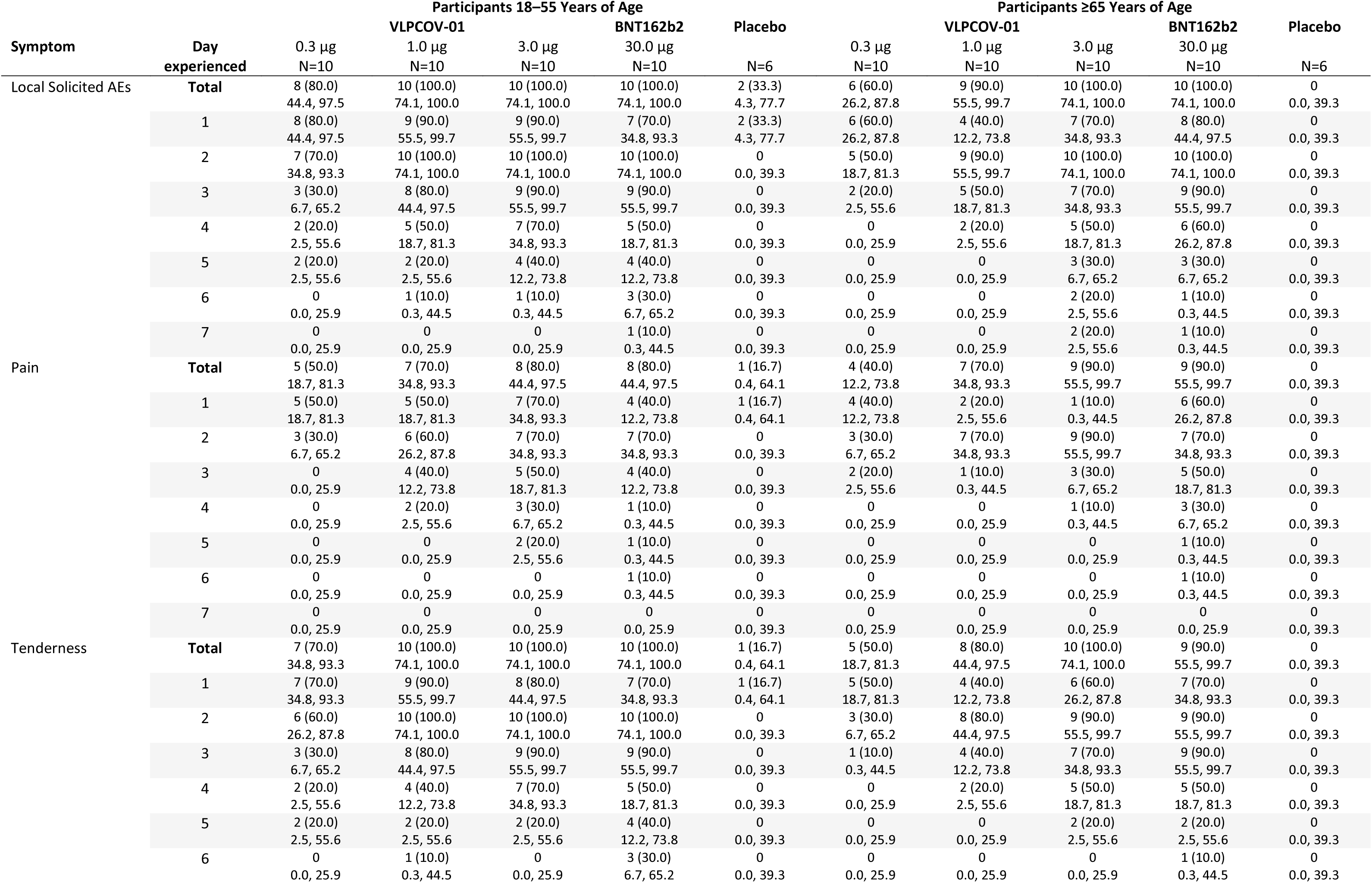
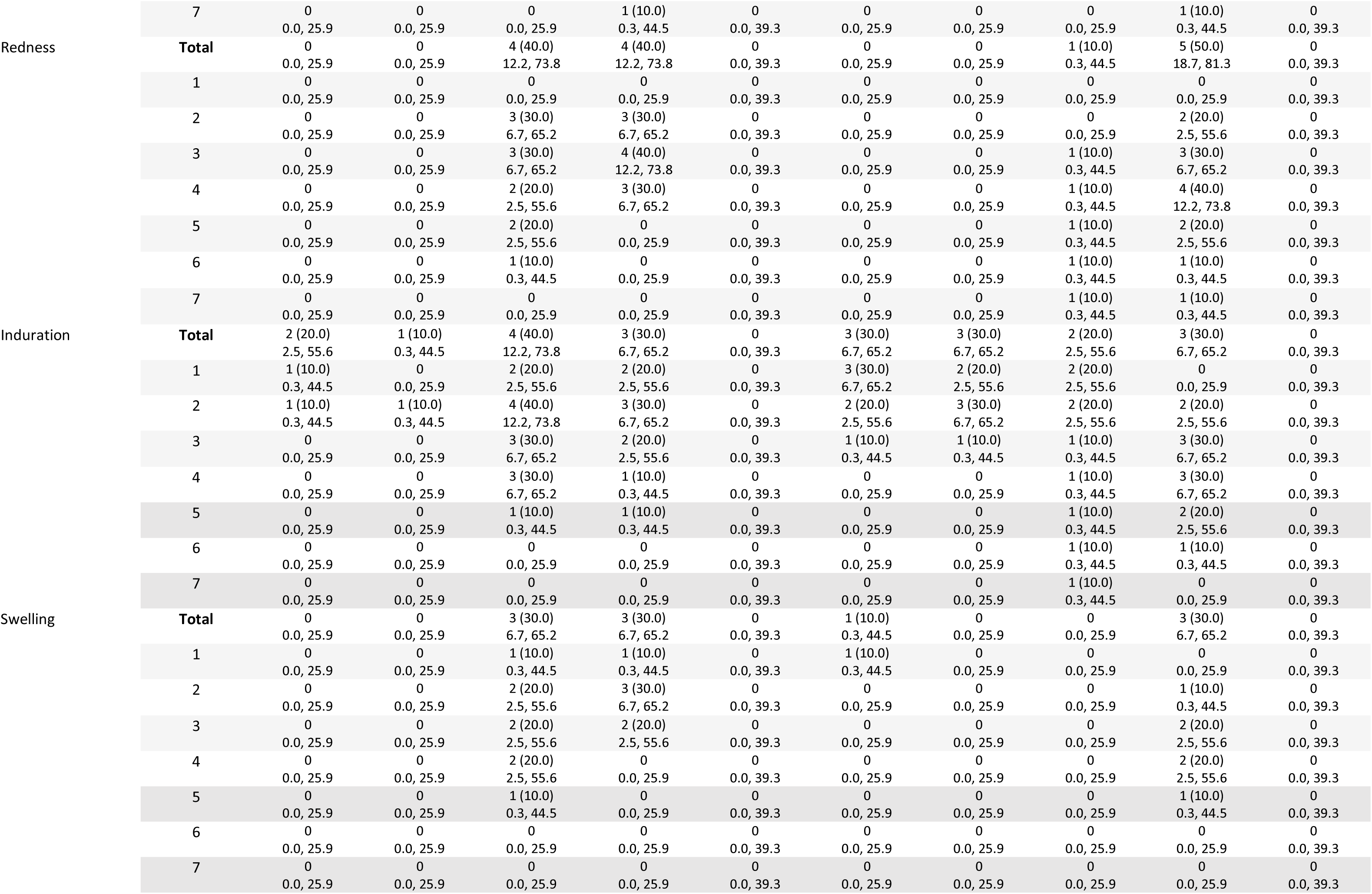
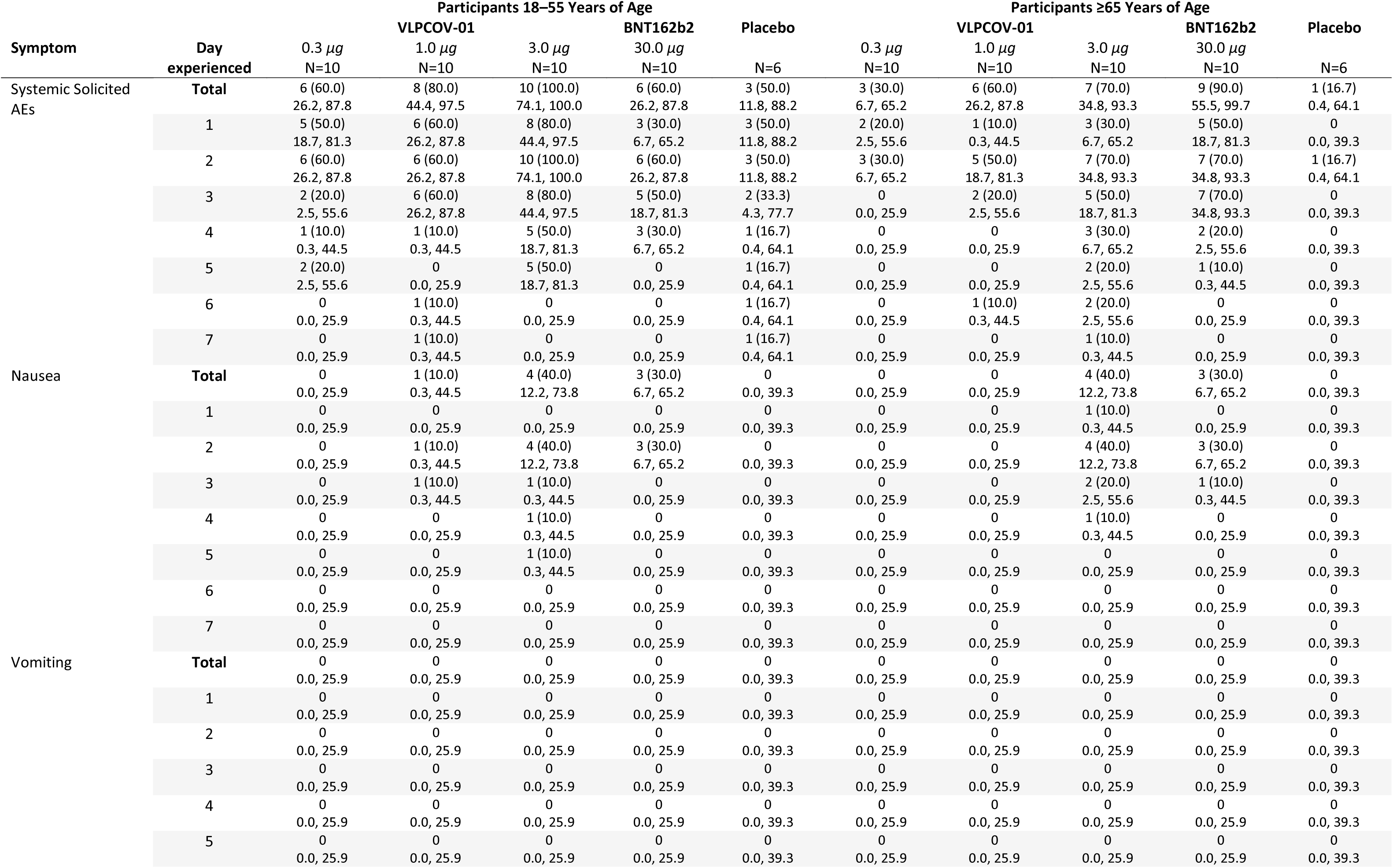
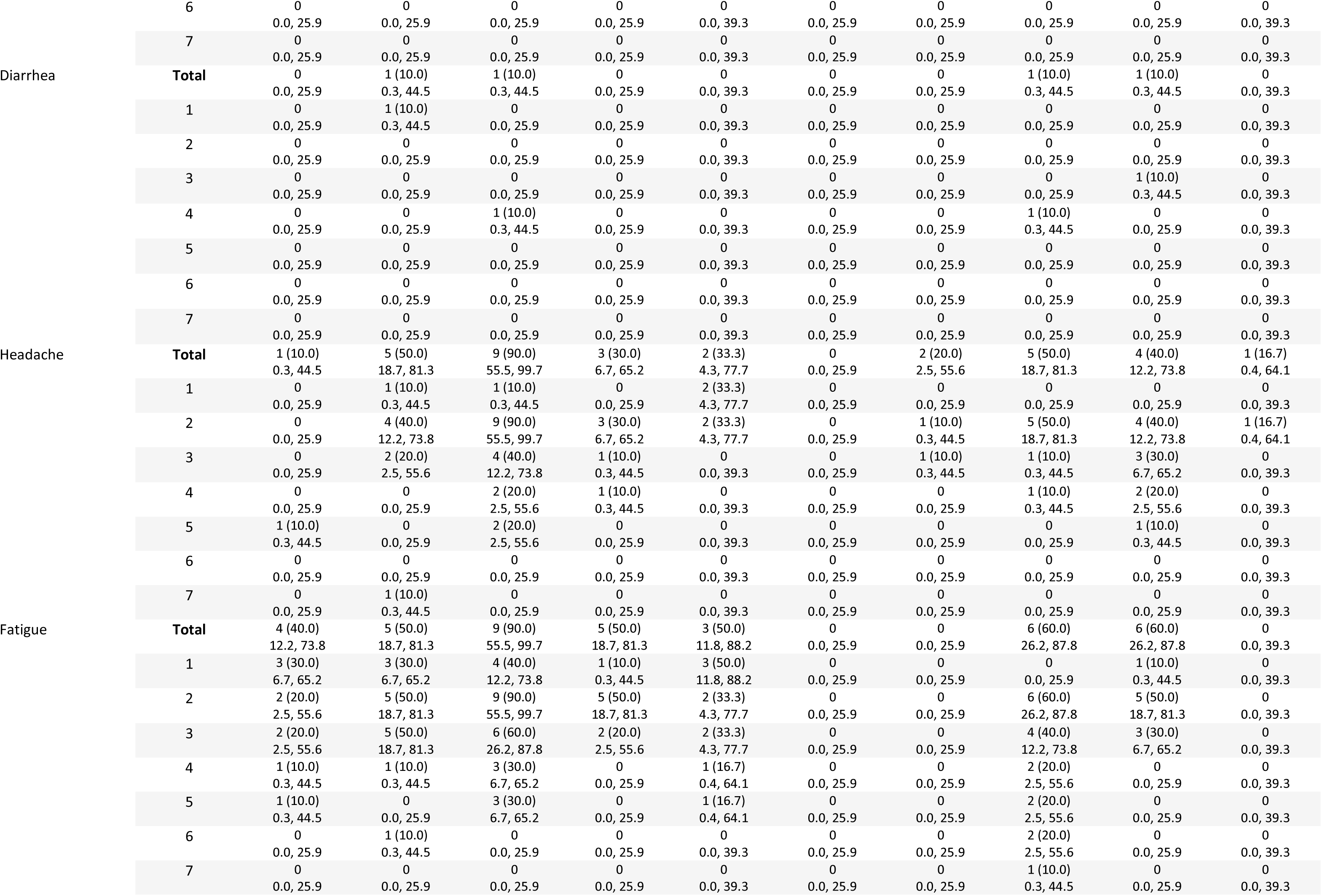
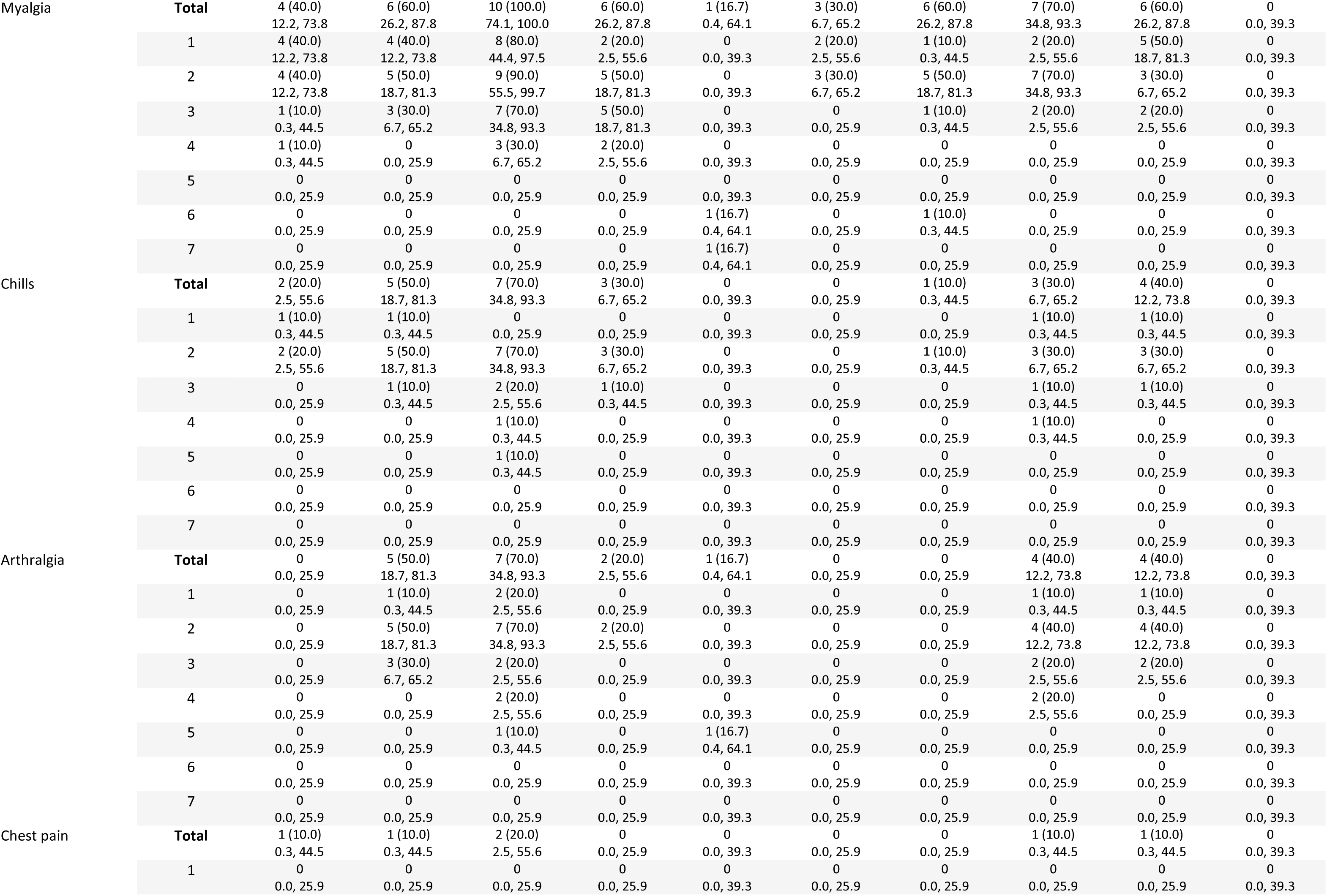
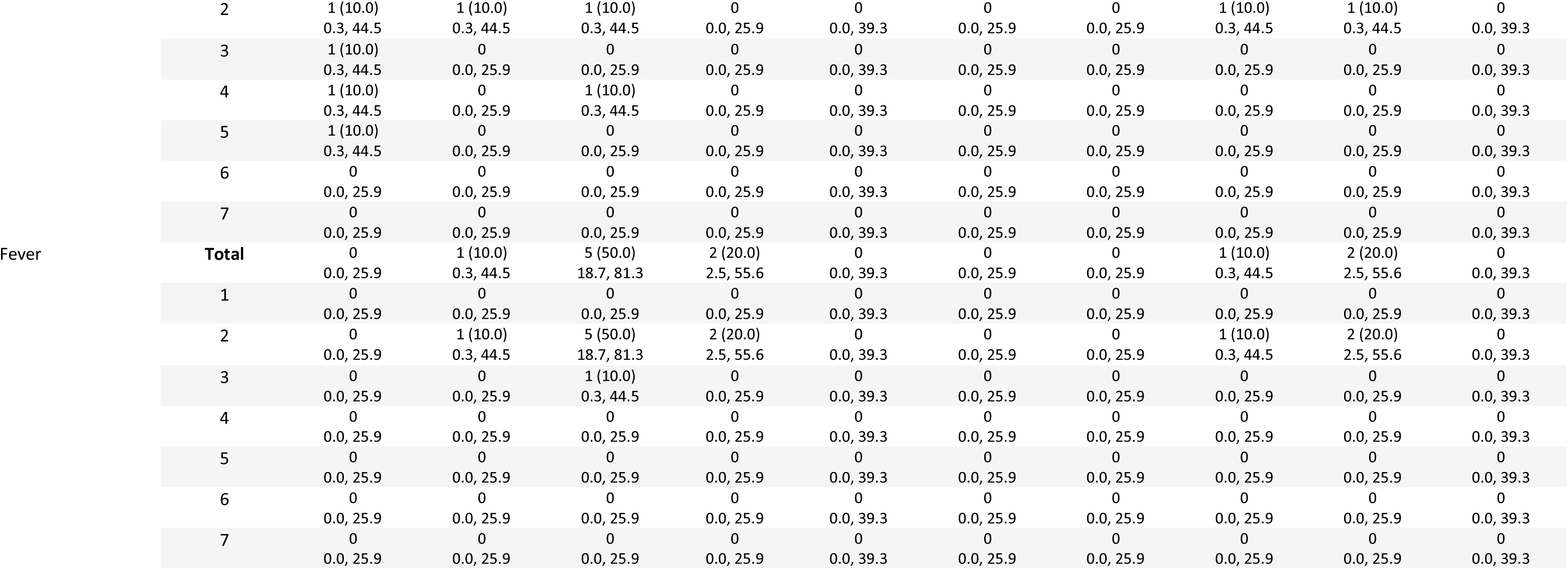
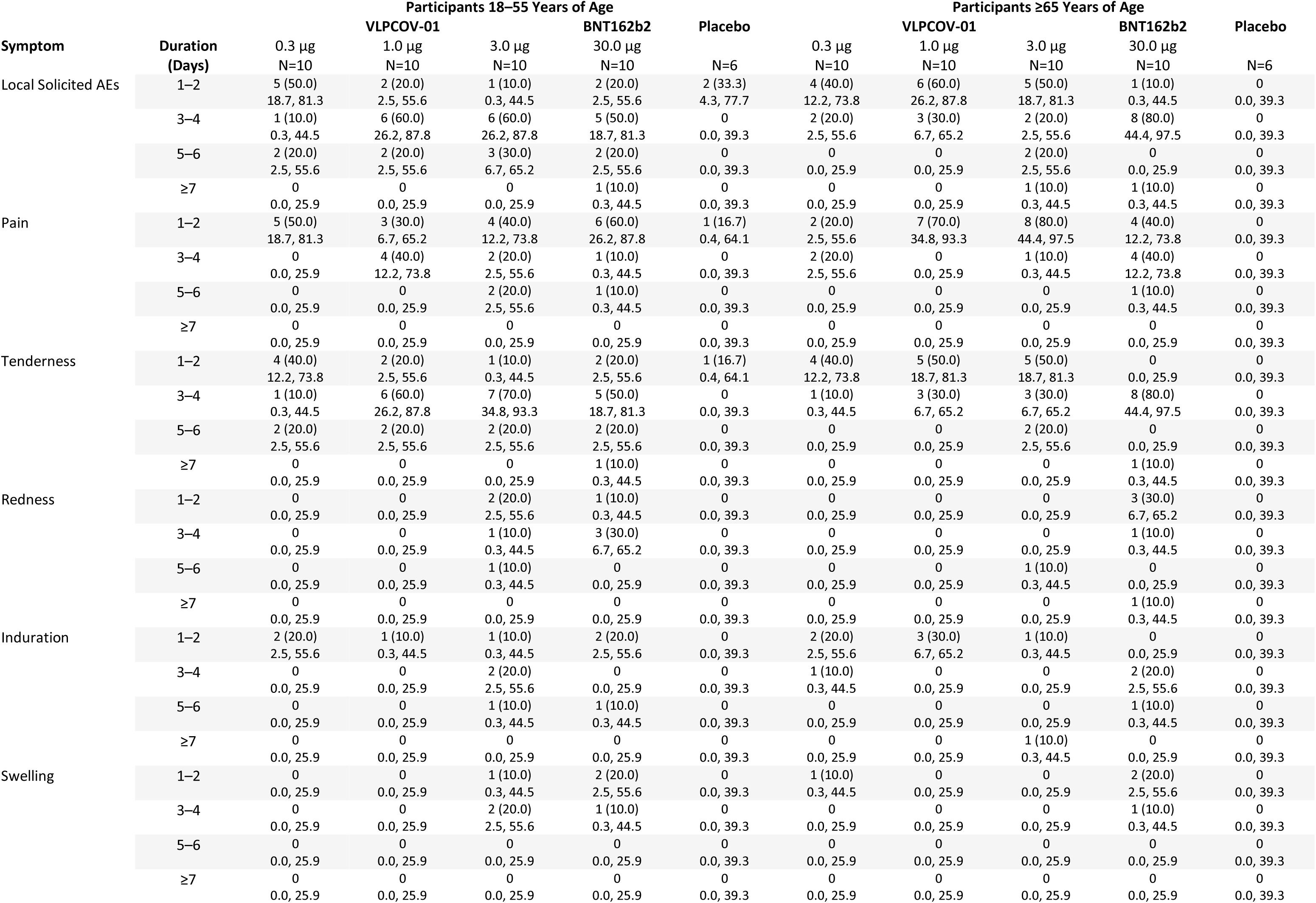
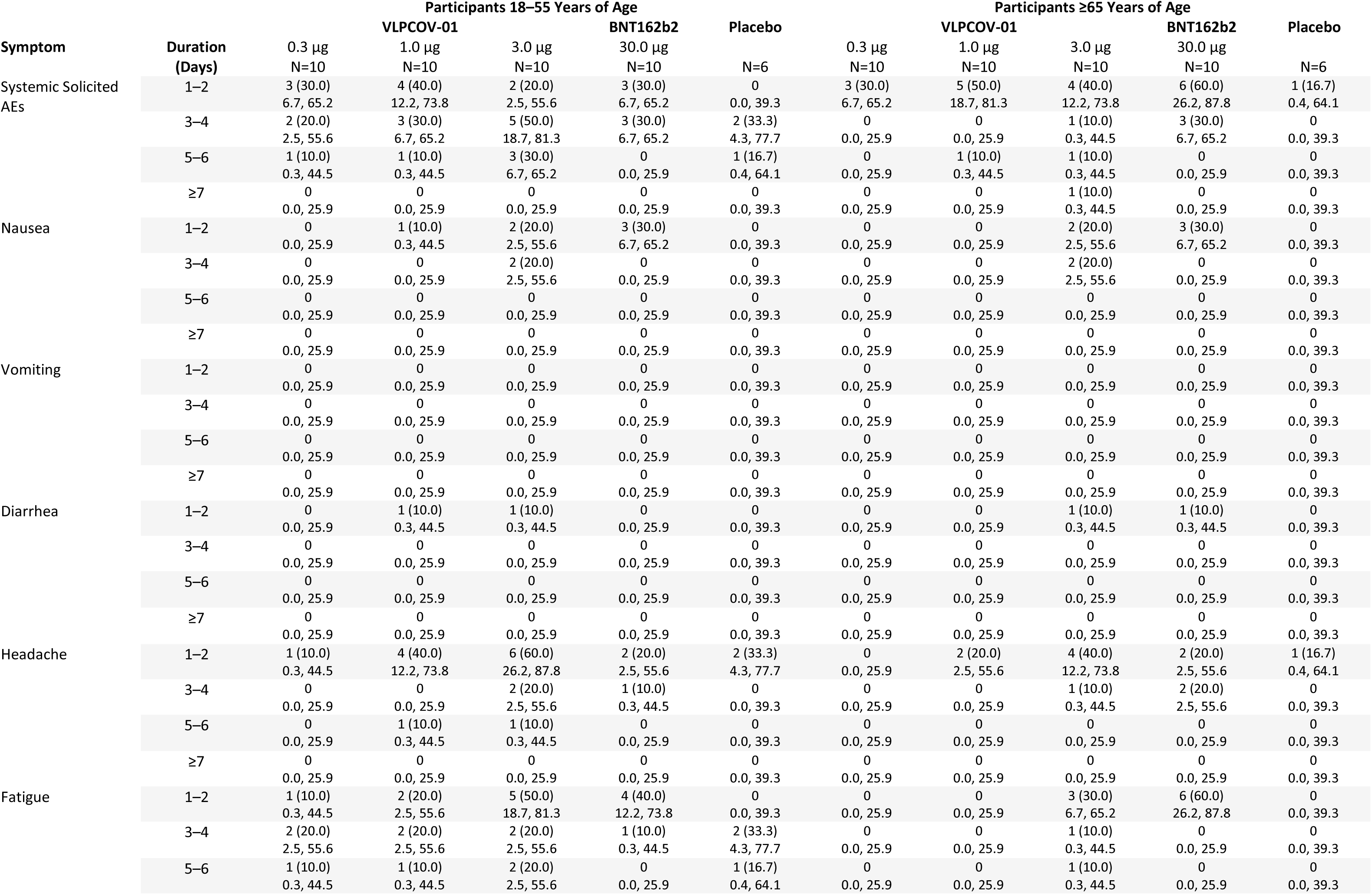
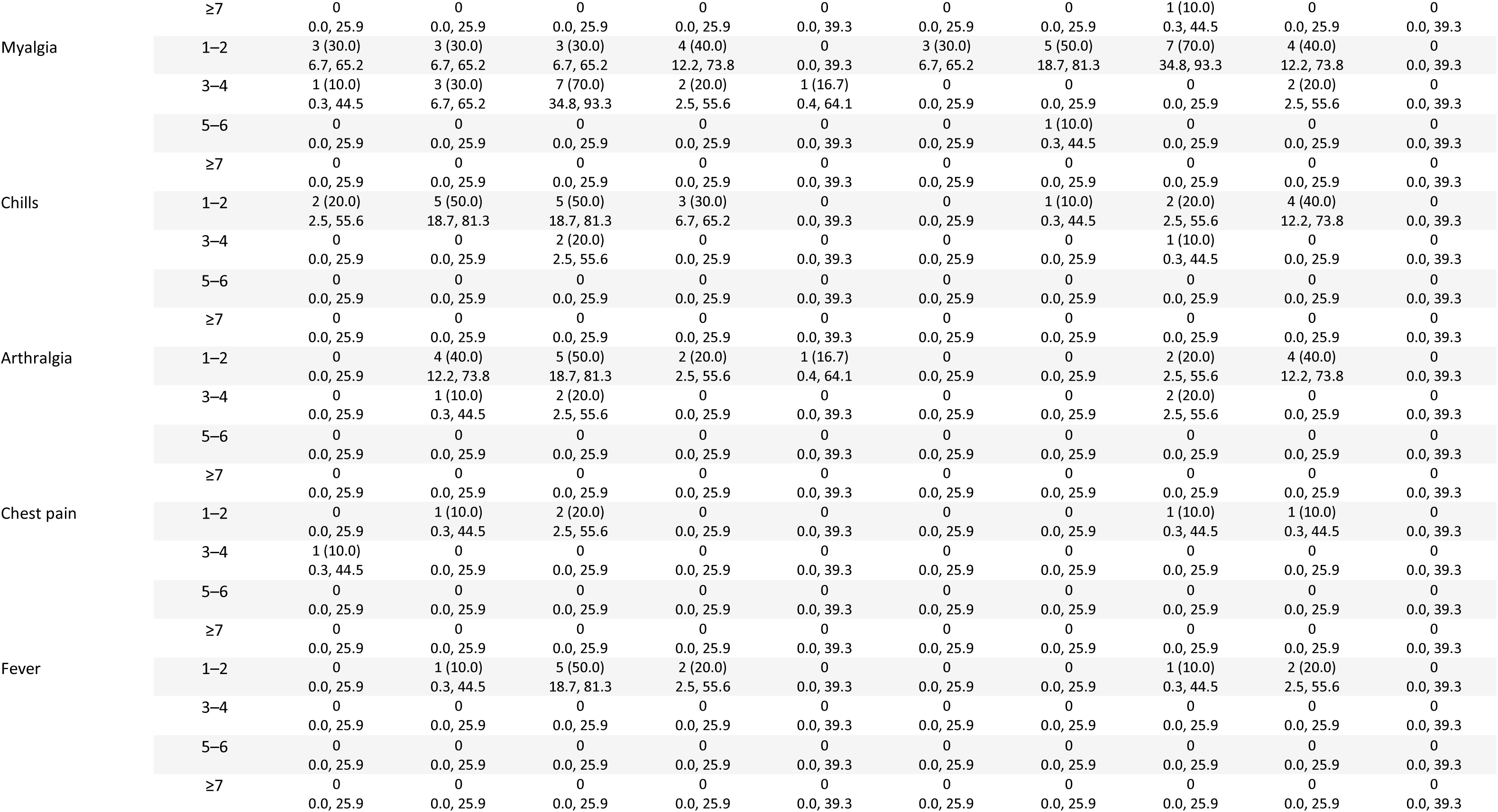
Onset and Duration of Solicited Adverse Events.

**Table S4.**
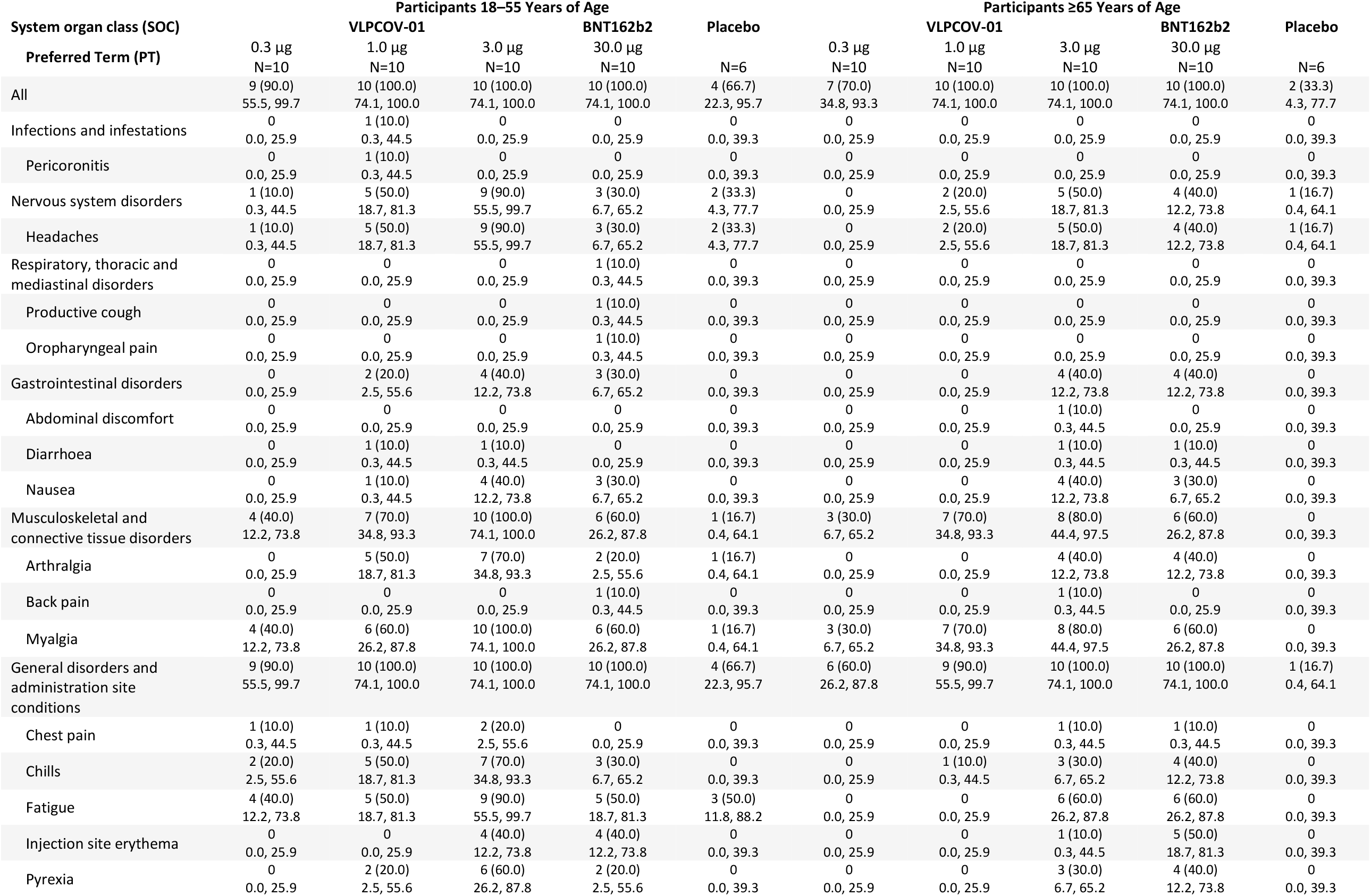
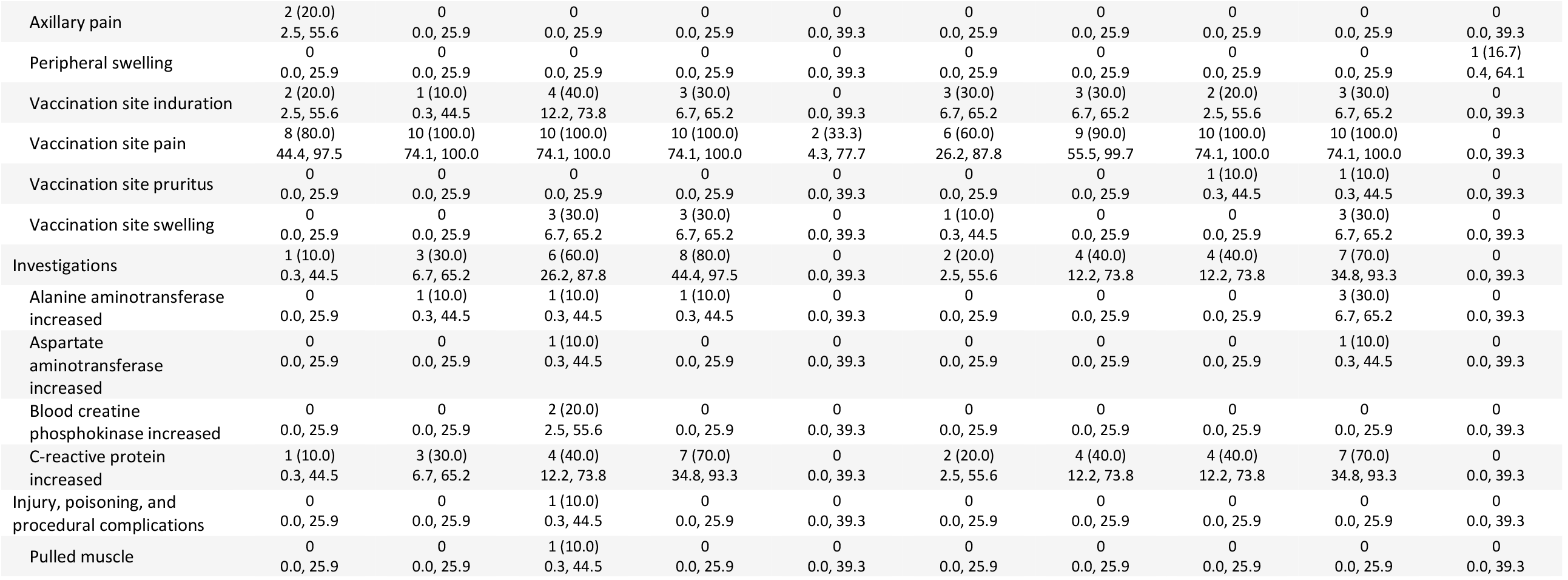
Unsolicited Adverse Events.

